# Mathematical assessment of the role of human behavior changes on SARS-CoV-2 transmission dynamics

**DOI:** 10.1101/2024.02.11.24302662

**Authors:** Binod Pant, Salman Safdar, Mauricio Santillana, Abba B. Gumel

## Abstract

The COVID-19 pandemic has not only presented a major global public health and socio-economic crisis, but has also significantly impacted human behavior towards adherence (or lack thereof) to public health intervention and mitigation measures implemented in communities worldwide. The dynamic nature of the pandemic has prompted extensive changes in individual and collective behaviors towards the pandemic. This study is based on the use of mathematical modeling approaches to assess the extent to which SARS-CoV-2 transmission dynamics is impacted by population-level changes of human behavior due to factors such as (a) the severity of transmission (such as disease-induced mortality and level of symptomatic transmission), (b) fatigue due to the implementation of mitigation interventions measures (e.g., lockdowns) over a long (extended) period of time, (c) social peer-pressure, among others. A novel behavior-epidemiology model, which takes the form of a deterministic system of nonlinear differential equations, is developed and fitted using observed cumulative SARS-CoV-2 mortality data during the first wave in the United States. Rigorous analysis of the model shows that its disease-free equilibrium is locally-asymptotically stable whenever a certain epidemiological threshold, known as the *control reproduction number* (denoted by *ℛ*_*C*_) is less than one, and the disease persists (i.e., causes significant outbreak or outbreaks) if the threshold exceeds one. The model fits the observed data, as well as makes a more accurate prediction of the observed daily SARS-CoV-2 mortality during the first wave (March 2020 -June 2020), in comparison to the equivalent model which does not explicitly account for changes in human behavior. Of the various metrics for human behavior changes during the pandemic considered in this study, it is shown that behavior changes due to the level of SARS-CoV-2 mortality and symptomatic transmission were more influential (while behavioral changes due to the level of fatigue to interventions in the community was of marginal impact). It is shown that an increase in the proportion of exposed individuals who become asymptomatically-infectious at the end of the exposed period (represented by a parameter *r*) can lead to an increase (decrease) in the control reproduction number (*ℛ*_*C*_) if the effective contact rate of asymptomatic individuals is higher (lower) than that of symptomatic individuals. The study identifies two threshold values of the parameter *r* that maximize the cumulative and daily SARS-CoV-2 mortality, respectively, during the first wave. Furthermore, it is shown that, as the value of the proportion *r* increases from 0 to 1, the rate at which susceptible non-adherent individuals change their behavior to strictly adhere to public health interventions decreases. Hence, this study suggests that, as more newly-infected individuals become asymptomatically-infectious, the level of positive behavior change, as well as disease severity, hospitalizations and disease-induced mortality in the community can be expected to significantly decrease (while new cases may rise, particularly if asymptomatic individuals have higher contact rate, in comparison to symptomatic individuals).

## Introduction

Throughout history, human civilization has repeatedly faced devastating disease pandemics, ranging from the bubonic plague (which caused 200 million deaths from 1347 to 1351), Smallpox (which caused 56 million deaths in 1520 alone), the plague of Justinian (which caused 30 to 50 million deaths from the year 541 to 542 AD), the third plague (which caused 12 million deaths in 1855 alone), the 1919 influenza pandemic (which claimed 40 million to 50 million lives), the HIV/AIDS epidemic (which resulted in 25 to 35 million deaths since its inception in 1981) [1–3] to the 2019 novel coronavirus pandemic (COVID-19). COVID-19, caused by the SARS-CoV-2, was first identified in the Wuhan province of China in December of 2019 and rapidly spread around the world causing the greatest public health challenge humans have faced since the 1918 influenza pandemic [1]. It has caused over 670 million confirmed cases and 7 million deaths globally during the first three years since its emergence [4]. In addition, it has induced severe socio-economic burden (with an estimated $12.5 trillion cost to the global economy projected through 2024 [5]). Further, Barber *et al*. [6] estimates that by November 14, 2021, about 44% of the world’s population was infected with COVID-19 at least once. Using a survey by the United States Census Bureau, the United States Centers for Disease and Prevention (CDC) estimated that one in five COVID-19-infected American adults has long COVID, a phenomenon described as COVID-19 “symptoms lasting three or more months after first contracting the virus” [7].

A pandemic of COVID-19’s devastating magnitude naturally invokes fear, unprecedented chaos, pandemonium, spread of mis(dis)information, mistrust, polarization resulting in both positive and negative behavior changes with respect to adherence (or lack thereof) of public health intervention and mitigation measures [8–13]. Numerous mathematical models, of varying types (such as compartmental, agents-based, social network, statistical, and machine learning models) have been developed and used in an attempt to study the impact of human behavior changes on the trajectory, transmission dynamics and overall burden of the pandemic. Specifically, some of these models have accounted for the behavior change due to the transmission of information through contacts between humans [14–18] or due to the prevalence of the pandemic [14, 15, 19]. For instance, using data collected through a contact diary-based survey, Kummer *et al*. [20] showed the impact of the seasonal change in the number of contacts made between individuals on the spread of the disease. Furthermore, d’Onofrio *et al*. used a deterministic compartmental model to study vaccination uptake behavior as a function of prevalence [21, 22]. Coelho and Codeco [23] used Bayesian inference to model vaccination behavior as a function of individual perception of vaccine safety [23]. Mooij *et al*. [24] developed a large-scale agent-based epidemic model that uses mobility data to calibrate the behavior of agents. Similarly, Del Valle *et al*. [22, 25] *used an agent-based model to showcase the impact of school closure and fear-based home isolation during a pandemic. Numerous studies have used network models to assess the impact of human behavior on disease spread and control [26, 27]*. *Finally, Frieswijk et al*. [28] developed a behavior-epidemic model where the classical *SIR* epidemic model is coupled with an evolutionary game-theoretic decision-making mechanism to incorporate self-protective measures taken by individuals during disease outbreaks.

Some studies have shown that a key factor that affects both COVID-19 transmission dynamics and human behavior changes with respect to the SARS-CoV-2 pandemic is the level of asymptomatic transmission in the community (i.e., disease transmission by infectious individuals who do not display clinical symptoms of the SARS-CoV-2 pandemic) [29–32]. Additionally, it is possible that for various reasons, such as repeated exposures and partial cross-immunity, the proportion of exposed individuals who become asymptomatic (as opposed to symptomatic) at the end of the exposed period may increase. Hence, it is important to assess the impact of the proportion of exposed individuals who become asymptomatic at the end of the exposed period on the spread of the disease and human behavior. Furthermore, numerous studies have shown that mathematical models for disease transmission that did not explicitly incorporate human behavior and heterogeneities failed to accurately capture the correct trajectory and burden of the pandemic [22, 33, 34]. For example, although some mathematical models have correctly captured the trajectory and burden of the ongoing COVID-19 pandemic [31, 35–37], numerous others that did not explicitly account for social and human behavior aspects have failed to correctly capture current and/or future course/trajectory of the pandemic (the agents-based model developed by the United Kingdom’s Scientific Advisory Group on Emergencies overestimated the burden of the SARS-CoV-2 Omicron at its peak by a factor of 20; and this discrepancy is attributed, in part, by the lack of explicit incorporation of human behavior elements into the model [38]).

The current study focuses on developing and using a novel mathematical model, which takes the form of a compartmental deterministic system of nonlinear differential equations, to assess the impact of human behavior changes on the transmission dynamics and control of infectious diseases. The proposed model specifically considers human behavior changes in two distinct population groups, one which strictly adheres to mitigation measures, and another one which ignores them. We assume that changes of human behavior in these two groups may occur due to a number of key (epidemiological) factors, such as (a) disease-related information received from members of the other group, (b) the level of symptomatic transmission in the community (c) proportion of non-symptomatic (susceptible, exposed, asymptomatic infectious and recovered) individuals in the community, (d) the level of publicly-available disease-induced mortality information and (e) fatigue to adherence to control and mitigation interventions in the community. Another notable feature of the model to be developed is the inclusion of the impact of asymptomatic transmission on the disease dynamics as well as on human behavior changes with respect to the spread and burden of the SARS-CoV-2 pandemic. The paper is organized as follows. The human behavior model is formulated in Section 2 with the functional form of behavior change functions derived in Section 2.1. A behavior-free model is also considered, which is a special case of the model with behavior changes. Both the behavior and behavior-free version of the model are fitted with observed data in Section 2.2. Numerical simulations are carried out in Section 3. The behavior change functions considered in this study are visualized in Section 3.1 and their impact on cumulative mortality and infection is also assessed through simulation. The impact of change in the proportion of exposed individuals who become asymptomatic on the reproduction number, mortality and behavior change is simulated in Section 3.3. Finally, the main results of this study are discussed and summarized in Section 4.

## 2 Formulation of the Mathematical Model

The SARS-CoV-2 transmission model that incorporates human behavior in the disease dynamics to be developed in this study is based on splitting the total human population at time *t*, denoted by *N* (*t*), into two groups based on adherence or lack thereof to public health interventions implemented to combat or mitigate the burden of the pandemic. Specifically, we define the following groups:

**Group 1:** Individuals who do not adhere to public health interventions and mitigation measures (also defined as the *non-adherent* group).

**Group 2:** Individuals who strictly adhere to public health interventions and mitigation measures (also defined as the *adherent* group).

The total population of individuals in group 1 at time *t*, denoted by *N*_1_(*t*), is subdivided into the mutually-exclusive compartments of non-adherent susceptible (*S*_1_(*t*)), exposed/latent (*E*_1_(*t*)), symptomatically-infectious (*I*_1_(*t*)), asymptomatically-infectious (*A*_1_(*t*)) and recovered (*R*_1_(*t*)) individuals, so that

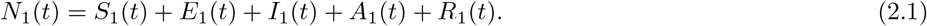

Similarly, the total population in group 2 at time *t*, denoted by *N*_2_(*t*), is sub-divided into the mutually-exclusive compartments of adherent susceptible (*S*_2_(*t*)), exposed/latent (*E*_2_(*t*)), symptomatically-infectious (*I*_2_(*t*)), asymptomatically-infectious (*A*_2_(*t*)) and recovered (*R*_2_(*t*)) individuals, so that

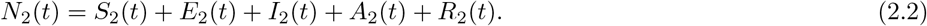

Thus, the total population at time *t, N* (*t*), is given by *N* (*t*) = *N*_1_(*t*) + *N*_2_(*t*). Some of the main behavior-related assumptions made in the formulation of the model are:

i. Individuals change their behavior during disease outbreaks based on the following epidemiological and social factors: (a) disease-related information members of one group received from members of the other group, (b) the level of symptomatic transmission in the community (c) the proportion of non-symptomatic (susceptible, exposed, asymptomatic infectious and recovered) individuals in the community, (d) the level of publicly-available disease-induced mortality information and (e) due to fatigue factor associated with adherence to interventions over a long-term period.
ii. Symptomatic individuals in group 2 do not change their behavior (i.e., they do not move to group 1) for the entire duration of their symptomatic status.
iii. The proportion of new recruited individuals into the community recruitment who are adherent to public health interventions depends on the proportion of symptomatic individuals in the community.

We define the following behavior-related transition rates:

a. 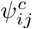: the rate at which individuals in group *i* change their behavior, and move to the other group *j* (with *i, j* = *{*1, 2*}* and *i ≠ j*), due to transmission of disease-related information received from contact with individuals in group *j*.
b. 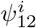: the rate at which individuals in group 1 positively change their non-adherent behavior and move to group 2 due to the level of symptomatic transmission in the community.
c. 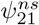: the rate at which individuals in group 2 negatively change their behavior and move to the non-adherent group 1 due to the proportion of non-symptomatic individuals in the community.
d. 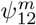: the rate at which individuals in group 1 positively change their behavior and move to the adherent group 2 due to the level of publicly-available disease-induced mortality information.
e. 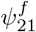: the rate at which individuals in group 2 negatively change their behavior and move to the non-adherent group 1 due to fatigue to adherence to intervention measures.

The functional forms of the aforementioned behavior-related rates will be derived in Section 2.1. In the formulation of the human behavior SARS-CoV-2 model, we let Π represent the recruitment rate of individuals into the population or community (and all recruited individuals are assumed to be susceptible). It is further assumed that a proportion, *p*, of these individuals strictly adhere to public health intervention and mitigation measures (i.e., *p*Π is the recruitment rate into the susceptible population of group 2), while the remaining proportion, 1− *p*, are assumed to not adhere to interventions (i.e., (1 − *p*)Π is the recruitment rate of individuals into the susceptible population in group 1). Individuals in all epidemiological compartments suffer natural death or removal at a rate *μ*. Individuals in group 1 (i.e., non-adherent individuals) acquire infection at a rate *λ* (*force of infection*), given by:

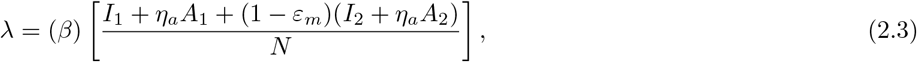

where *β* is the effective contact rate associated with disease transmission by symptomatically-infectious individuals, *η*_*a*_ is a modification parameter accounting for the variability in disease transmission by asymptomatically-infectious individuals, in comparison to symptomatically-infectious individuals and 0 ≤ *ε*_*m*_ *<* 1 is the *outward* efficacy of the public health intervention and mitigation measures (e.g., face mask) to prevent the transmission of infection by infectious adherent individuals (group to susceptible individuals [31, 39]. For simplicity, it is assumed that the *inward* efficacy of the public health intervention measures to prevent susceptible individuals in group from acquiring infection, following contact with infectious individuals, is the same as the outward efficacy, and denoted by *ε*_*m*_.

Individuals in group 1 (non-adherents) positively change their behavior and become adherents due to the positive information they received about the disease from individuals in group 2 (due to contact at a rate 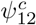). They also change their behavior positively and move to group 2 due to the level of symptomatic transmission in the community (at a rate 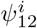) and due to the level of disease-induced mortality in the community, obtained from publicly-available sources (at a rate 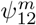). Similarly, individuals in group 2 negatively change their behavior and move to group 1 due to: the contact-related information received from individuals in group 1 (at a rate *ψ*^*c*^), the proportion of non-symptomatic individuals in the community (at a rate 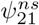) and due to fatigue to adherence of intervention (at a rate 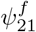). Adherents susceptible individuals acquire infection at a reduced rate (1 − *ε*_*m*_)*λ*, where 0 *< ε*_*m*_ *<* 1 is the efficacy of public health intervention and mitigation measures implemented in the community to prevent the acquisition of infection. Let *σ* represents the rate at which exposed (i.e., newly-infected) individuals progress to to either the asymptomatically-infectious class (at a rate *rσ*, where 0 ≤ *r*≤ 1 is the proportion of these individuals that become asymptomatically-infectious at the end of the exposed period) or develop clinical symptoms of the disease (at a rate (1 − *r*)*σ*, where the complement, 1− *r*, is the proportion of exposed individuals who display clinical symptoms of the disease at the end of the exposed period). Symptomatic (asymptomatic) infectious individuals are assumed to recover at a rate *γ*_*i*_(*γ*_*a*_), and symptomatic-infectious individuals in group 1 (group 2) suffer disease-induced mortality at a rate *δ*_1_(*δ*_2_). It follows, based on the above assumptions and derivations, that the endemic model for the SARS-CoV-2 pandemic, that explicitly incorporates elements of human behavior during the outbreaks of the disease, is given by the following deterministic system of nonlinear differential equations (where a dot represents differentiation with respect to time *t*):

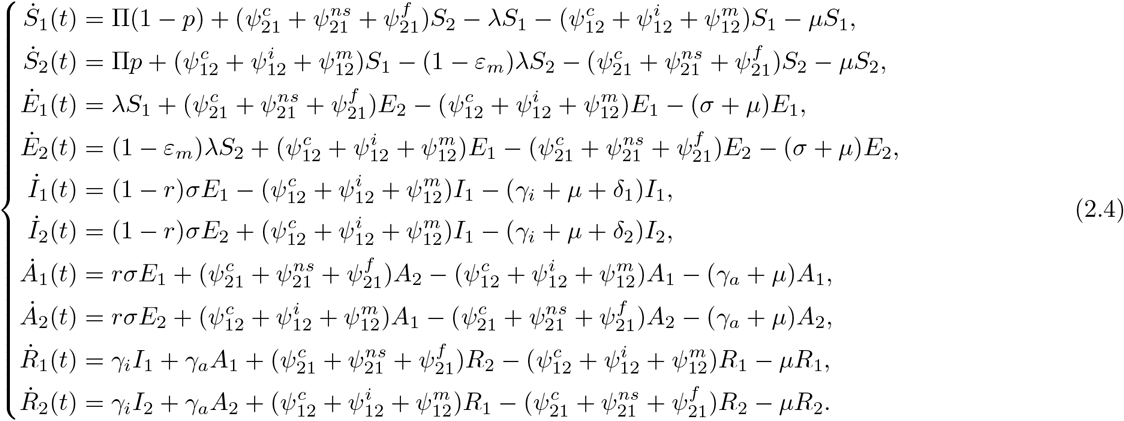

In the behavior-epidemiology model (2.4), the proportion *p* of individuals recruited into the community who adhere to public health interventions is assumed to defend on the proportion of individuals who are symptomatic at time *t*, and is defined as:

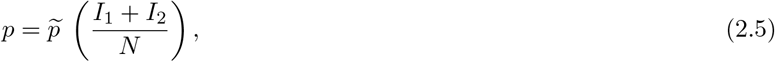

where 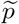 is the maximum proportion of recruited individuals who are adherent. The proportion *p* is behavior-related, since it is a function of the total proportion of symptomatic individuals in the community. It is zero if there are no symptomatic individuals in the community, and it rises to its maximum 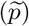 if the proportion of symptomatic individuals in the community tend towards one; in other words, newly-recruited individuals choose to remain in group 1 if the proportion of symptomatic individuals in the community is small, and opt to move to group 2 if the proportion of symptomatic individuals is large).

It is convenient to define the rate of change of cumulative mortality due to the disease as

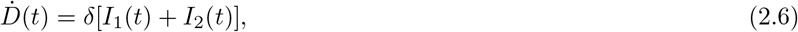

from which it follows that the daily mortality on day *k* (denoted by *M*_*k*_) is given by:

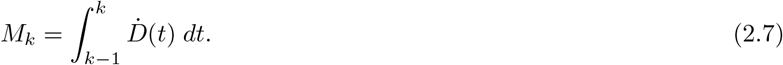

Figure 1 depicts the flow diagram of the model, and the state variables and parameters are described in Tables 1 and 2, respectively.

**Table 1:**
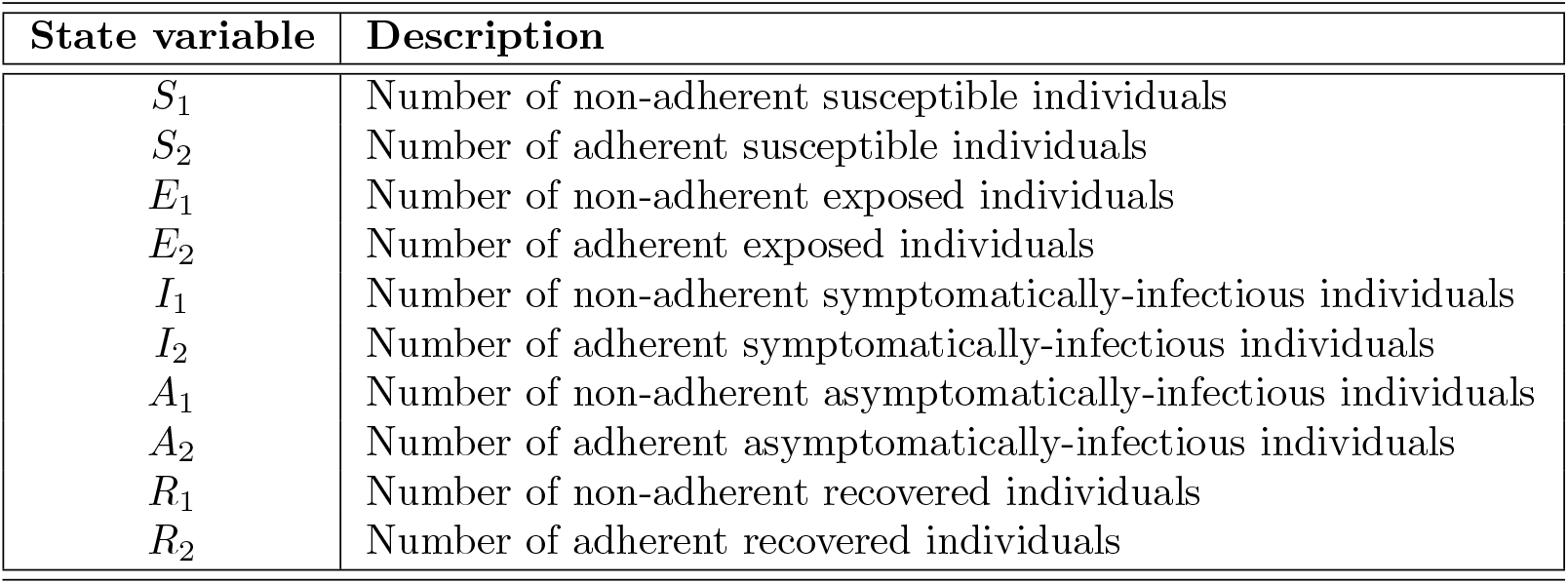
Description of the state variables of the behavior-epidemiology model (2.4).

**Table 2:** Description of the parameters of the model (2.4).

| Parameter | Description |
| --- | --- |
| $p$ | Proportion of new recruitment that is adherent |
| $\Pi(1 - p)$ | Recruitment rate into non-adherent susceptible population |
| $\Pi p$ | Recruitment rate into adherent susceptible population |
| $\mu$ | Natural death rate |
| $\varepsilon_m$ | Average efficacy of an intervention (such as masking or social distancing) |
| $\beta$ | Effective contact rate associated with individuals in $I_1$ compartment |
| $\eta_a$ | Modification parameter for the infectiousness of the individuals in asymptomatic class |
| $r$ | Proportion of exposed individuals who become asymptotically-infectious at the end of the exposed period |
| $1 - r$ | Proportion of exposed individuals who show clinical symptoms of the disease at the end of the exposed period |
| $r\sigma$ | Progression rate from exposed to asymptotically-infectious class |
| $(1 - r)\sigma$ | Progression rate from exposed to symptomatically-infectious class |
| $\gamma_i(\gamma_a)$ | Recovery rate for individuals in the symptomatically- (asymptotically-) infectious class |
| $\delta_1$ | Disease-induced mortality rate for individuals in the $I_1$ compartment |
| $\delta_2$ | Disease-induced mortality rate for individuals in the $I_2$ compartment |
| $\psi_{12}^c$ | Rate at which non-adherent individuals become adherent due to contact with adherents |
| $\psi_{21}^c$ | Rate at which adherent individuals become non-adherent due to contact with non-adherents |
| $\psi_{12}^i$ | Rate at which non-adherent individuals become adherent due to the level of symptomatic transmission in the community |
| $\psi_{21}^{ns}$ | Rate at which adherent individuals become non-adherent due to the proportion of non-symptomatic individuals in the community |
| $\psi_{12}^m$ | Rate at which non-adherent individuals become adherent due to disease-induced mortality |
| $\psi_{21}^f$ | Rate at which adherent individuals become non-adherent due to high fatigue level |
| $\alpha_{12}^c$ | Maximum rate of behavior change of non-adherent individuals due to contact with adherent individuals |
| $q_{12}^c$ | Probability of influence of non-adherent individuals due to contact with adherent individuals |
| $\alpha_{21}^c$ | Maximum rate of behavior change of adherent individuals due to contact with non-adherent individuals |
| $q_{21}^c$ | Probability of influence of adherent individuals due to contact with non-adherent individuals |
| $\alpha_{12}^i$ | Maximum rate of behavior change of non-adherent individuals due to the level of symptomatic transmission |
| $q_{12}^i$ | Probability of influence of non-adherent individuals due to the level of symptomatic transmission |
| $\alpha_{21}^{ns}$ | Maximum rate of behavior change of adherent individuals due to the proportion of non-symptomatic transmission in the community |
| $q_{21}^{ns}$ | Probability of influence of adherent individuals due to the proportion of non-symptomatic individuals |
| $\alpha_{12}^m$ | Maximum rate of behavior change of non-adherent individuals due to disease-induced mortality |
| $q_{12}^m$ | probability of influence of non-adherent individuals due to disease-induced mortality |
| $\alpha_{21}^f$ | Maximum rate of behavior change of adherent individuals due to high fatigue level to interventions |
| $q_{21}^f$ | Probability of influence of adherent individuals due to high fatigue level to interventions |
| $t_f$ | Threshold time after which adherent individuals begin to experience pandemic fatigue |
| $1/\zeta$ | The number of reported disease-induced death above which behavior change due to mortality rapidly increases |
| $K$ | Half saturation constant |

**Figure 1:**
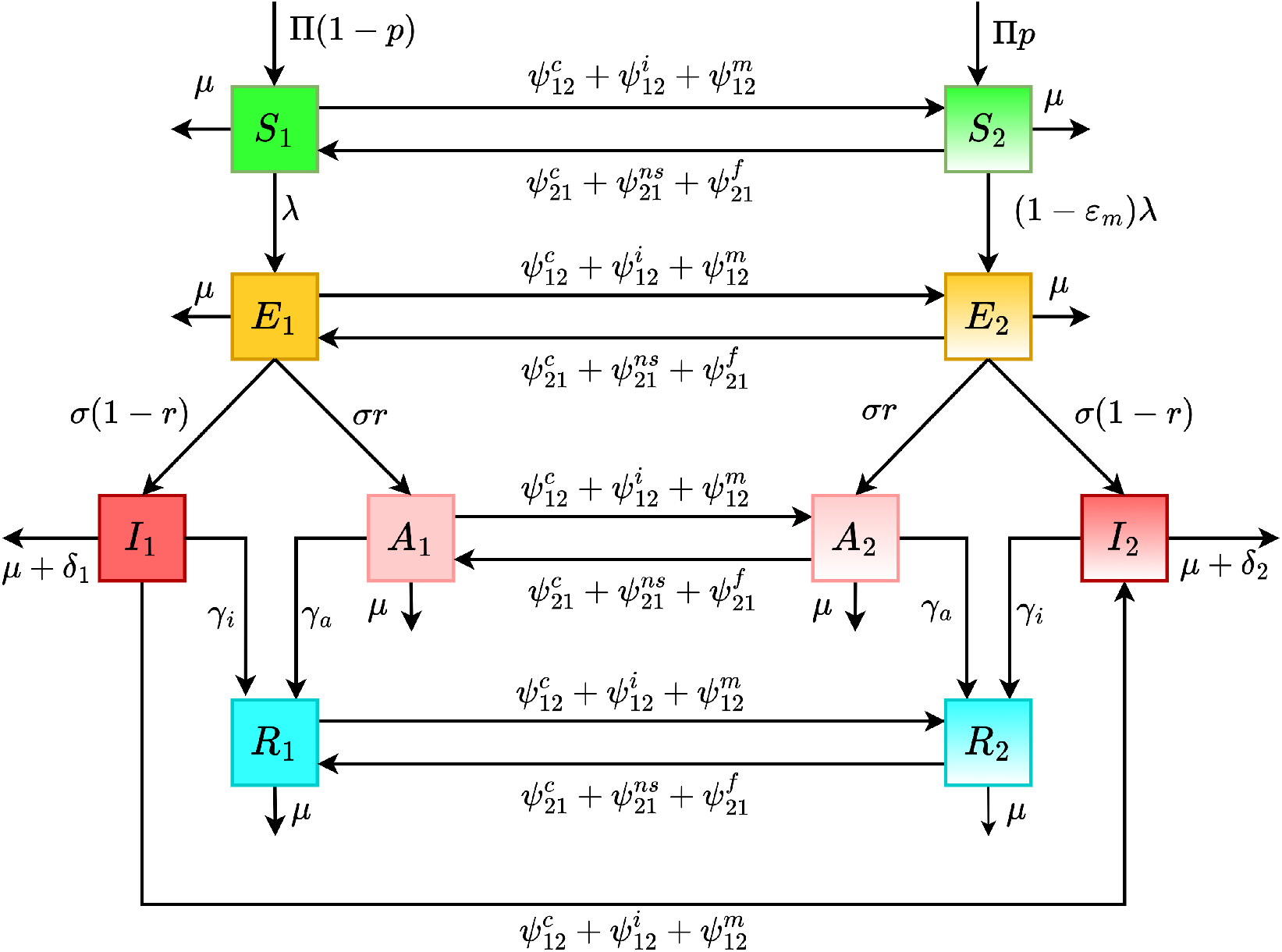
Flow diagram of the behavior-epidemiology model (2.4), where *λ* and *p* are defined in Equations (2.3) and (2.5), respectively. The state variables and parameters of the model are described in Tables 1 and 2, respectively.

### 2.1 Derivation of the functional forms of behavior-related parameters of the model

The behavior-related transition rates of the behavior-epidemiology model (2.4) are described and derived below.

#### 2.1.1 Behavior change due to information received from contact with members of the other group 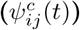

Since the behavior-epidemiology model to be developed in this study stratifies the total population into two subgroups, namely group 1 of individuals who do not adhere to public health control and mitigation interventions and group 2 consisting of individuals who strictly adhere to such interventions, contacts between individuals of one group with those of the other could induce behavior change with respect to the adherence (or lack thereof) to interventions (due, for instance, to *peer influence or pressure*). It is intuitive to assume that the probability of such contact-induced behavior change depends on the capacity of (or degree or extent to which) members of one group to influence members of the other and the relative size of the other group [14–18]. Recall that 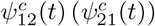 represents the rate at which non-adherent (adherent) individuals become adherents (non-adherents), at time *t*, following contacts with adherent (non-adherent) individuals in the community. The rate 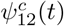can then be defined as the product of the maximum rate of behavior change of non-adherent individuals to become adherents due to contacts with adherent individuals (denoted by 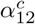), the probability of influence adherent individuals have on non-adherent individuals to become adherent due to transmission of disease-related information by contact (denoted by 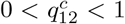 and the current proportion of adherent individuals in the community 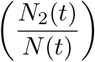. Thus, the rate 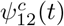 is given by:

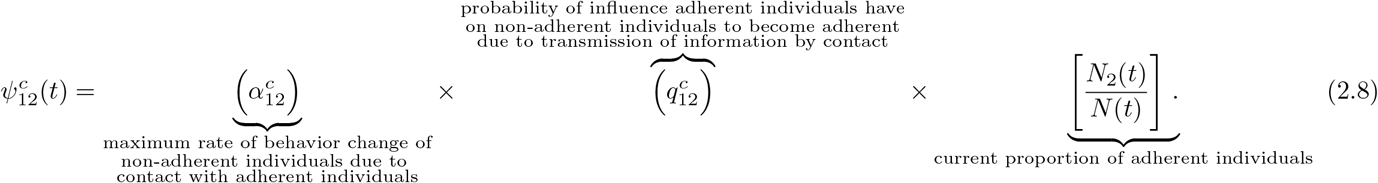

Similarly, the rate at which adherent individuals in the community become non-adherent due to contacts with non-adherent individuals at time 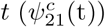 is defined as the product of the maximum rate of behavior change of adherent individuals due to contacts with non-adherent individuals (denoted by 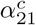), the probability of influence non-adherent individuals have on adherent individuals to become non-adherents due to transmission of disease-related information by contacts (denoted by 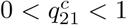 and the current proportion of non-adherent individuals in the community 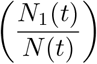, so that:

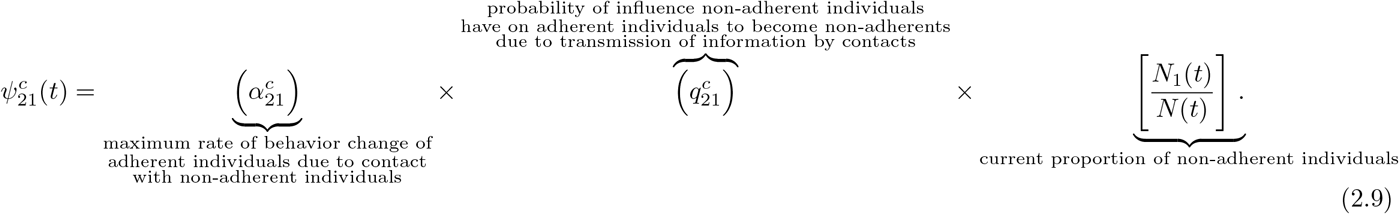

#### 2.1.3 Behavior change due to the level of symptomatic transmission 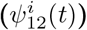 in community

Individuals can also change their behavior with respect to adherence to interventions based on the current relative level of symptomatic transmission in the community [14]. Let 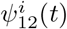 represent the rate at which non-adherent individuals become adherent due to the level of symptomatic transmission in the community. The rate 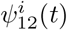 is defined as the product of the maximum behavior change of non-adherent individuals due to the level of symptomatic transmission in the community 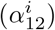, the probability of influence on non-adherent individuals due to the level of symptomatic transmission in the community 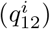 and the relative level of symptomatic transmission in the community (given by the Holling Type-II saturation incidence function (*I*_1_(*t*) + *I*_2_(*t*))*/*(*K* + *I*_1_(*t*) + *I*_2_(*t*)), where *K >* 0 is the saturation constant). That is,

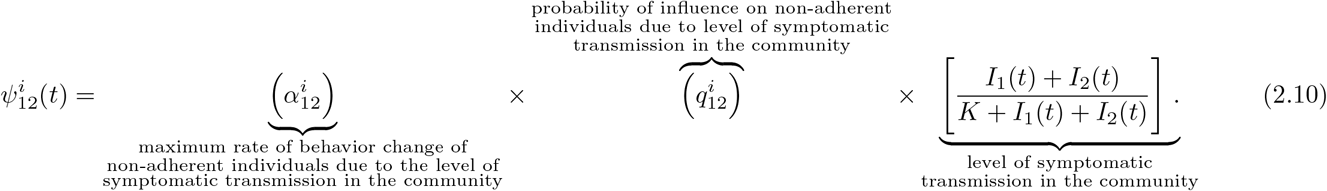

#### 2.1.3 Behavior change due to the proportion of non-symptomatic 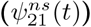 pool in community

Individuals can also change their behavior due to the proportion of non-symptomatic (i.e., susceptible, exposed, asymptomatic, and recovered) individuals in the community [15]. Let 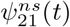 represent the rate at which adherent individuals become non-adherent due to the current proportion of non-symptomatic individuals in the community. The parameter for the rate of behavior change of adherent individuals to become non-adherent due to the proportion of non-symptomatic individuals in the community 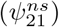 is given by the product of the maximum rate of behavior change of adherent individuals due to the proportion of non-symptomatic individuals in the community 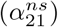, the probability of influence on adherent individuals due to the current size of the non-symptomatic pool in the community 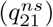 and the proportion of the non-symptomatic pool in the community (given by (*S*_1_(*t*) + *S*_2_(*t*) + *E*_1_(*t*) + *E*_2_(*t*) + *A*_1_(*t*) + *A*_2_(*t*) + *R*_1_(*t*) + *R*_2_(*t*))*/N* (*t*)). Hence,

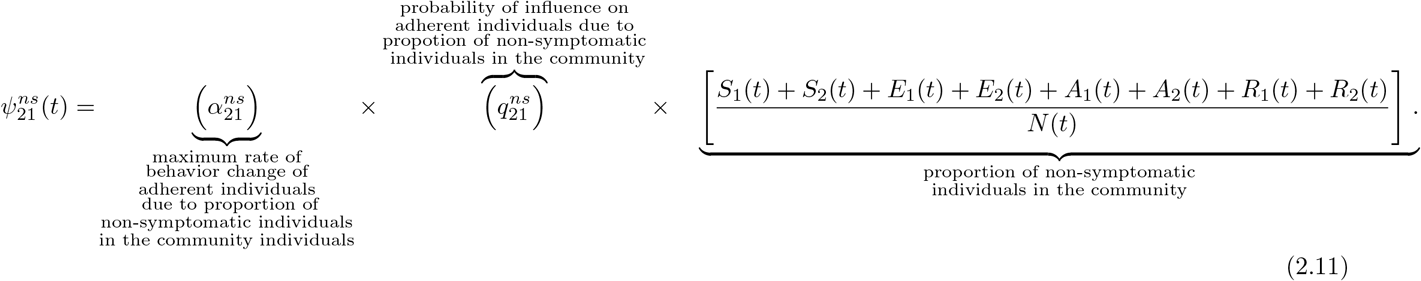

#### 2.1.4 Behavior change due to level of daily reported disease-induced mortality 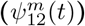

Individuals can also change their behavior due to the level of daily reported disease-induced mortality (obtained from publicly-available information sources, such as the print/audio/visual/social and the internet) [14]. To model the rate of behavior change due to such access to information on the level of daily disease-induced mortality in the community, we first assume that the level of disease-induced mortality in the community only causes behavior change in group 1, and not in group 2 (i.e., only the non-adherent individuals can change their behavior to become adherent in fear of succumbing to the disease; whereas those that are strictly adherent, in group 2, are not expected to negatively change their behavior, when daily disease-induced mortality is on the rise, and move to group 1). The rate 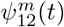 can then be expressed as a product of the maximum rate of behavior change of non-adherent individuals due to the level of daily reported disease-induced mortality in the community (denoted by 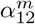), the probability of influence adherent individuals have on non-adherent individuals due to the size of the reported daily disease-induced mortality in the community (denoted by 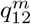) and the current level of disease-induced mortality (modeled by the Ivlev function [40], (1 − *e*^−*ζM* (*t*)^), where *M* (*t*) is the daily mortality on day *t* of the pandemic, as defined by Equation (2.7) and 1*/ζ*(with 0 *< ζ*≤ 1) could be thought of as the threshold number of reported daily disease-induced mortality above which fear or *panic wave* begin to rapidly spread in the community [14]). Thus,

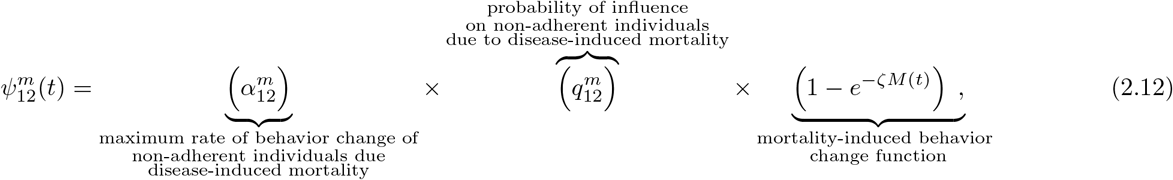

#### 2.1.5 Behavior change due to high level of fatigue to interventions in the community 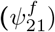

Behavior change “requires a person to disrupt a current habit while simultaneously fostering a new, possibly unfamiliar, set of actions” [41]. Thus, perhaps for that reason, behavior change, even if it is triggered by health-related reasons (i.e., triggered by the need to adhere to public health intervention and mitigation measures to minimize the risk of acquiring infection, severe disease, hospitalization or even death), can often be quite difficult to sustain. To model behavior change due to fatigue to adherence to public health intervention and mitigation measures, it is plausible to, first of all, assume that only individuals in group 2 (the adherent group) can develop intervention fatigue (due to being strictly adherent for an extended period of time) and potentially negatively change their behavior to become non-adherents. For simplicity, it is assumed that adherent individuals begin to experience fatigue of adherence to intervention after a certain fatigue time threshold (denoted by *t*_*f*_) has been met. This factor can be modeled using a Heaviside-like function (*H*^*f*^), given by:

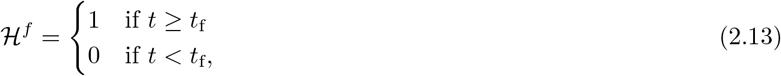

with *H*^*f*^ set to 1 if the fatigue time threshold has been met or exceeded (i.e., *t* ≥ *t*_*f*_) and *H*^*f*^ = 0 if the threshold has not been met, and adherents are not experiencing intervention fatigue (i.e., *t < t*_*f*_). The rate at which adherent individuals become non-adherent due to fatigue 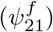 can then be expressed as a product of the maximum rate of behavior change of adherent individuals due to fatigue (denoted by 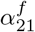), the probability of influence non-adherent individuals have on adherent individuals due to high fatigue level to intervention (denoted by 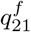) and the fatigue threshold time (given by the function ℋ^*f*^).

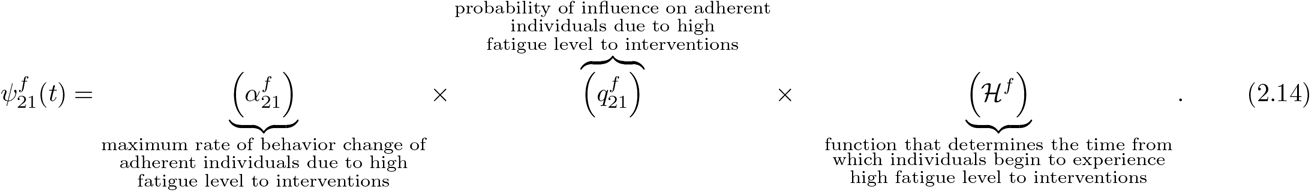

Based on the above assumptions and derivations, the behavior-epidemiology model for the transmission dynamics of the SARS-CoV-2 pandemic in a community (that incorporates elements of human behavior changes in response to the pandemic) is given by the following deterministic system of nonlinear differential equations (the flow diagram of the model is depicted in Figure 1, and the variables and parameters of the model are described in Tables 1 and 2, respectively):

### 2.2 Data-fitting and parameter estimation of behavior-epidemiology model (2.4)

The behavior-epidemiology model (2.4) consists of many parameters, the values of many of which (at least 16) are known from the literature. The values of a few of the parameters, particularly the disease-induced mortality rate (*δ*_1_ and *δ*_2_; although, for simplicity, we assume that *δ* = *δ*_1_ = *δ*_2_ since it is reasonable to assume that both adherent and non-adherent infected individuals die at the same rate due to, for example having access to the same quality of disease-induced mortality prevention treatment), the effective contact rate of symptomatic individuals (*β*), the modification parameter for the effective contact rate of asymptomatic individuals (*η*_*a*_) and the product of the maximum rate of behavior change from one group to another and the associated parameter of influence 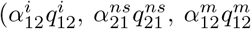 and 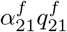 are unknown and will be estimated through fitting. Furthermore, behavior-related functions are computed using the fixed and fitted parameters as well as the associated state variables at time *t*. The model (2.4) will be fitted and cross-validated using the cumulative mortality data for the COVID-19 pandemic in the United States for the period from March 1st, 2020 to June 18th, 2020, which corresponds to the first wave of the pandemic in the United States. Specifically, the model will be fitted using a segment of this data, for the period between March 1, 2020, to May 29, 2020, and the remaining data is used to cross-validate the model. Additionally, some of the initial conditions will be fitted from the data as well.

Before describing the data fitting of the behavior-epidemiology, the values of the 16 known parameters (also known as *fixed parameters*) of the model are described below.

#### 2.2.1 Values of the fixed (known) parameters of the behavior-epidemiology model

The values of the 16 known parameters of the model (2.4) (tabulated in Table 3) are described as follows: the value of the daily recruitment rate parameter (Π) is obtained from using the census data for the United States, and noting that the total population of the United States prior to the emergence of the SARS-CoV-2 pandemic (i.e., at the disease-free equilibrium) is Π*/μ* (where 1*/μ* is the average lifespan). Since the total population of the United States was approximately 331.4 million [42] and the average lifespan is 77.8 years [43] (so that 1*/μ* = 77.8 years; hence, *μ* = 3.52 *×* 10^−5^ *per* day), it follows that Π = 331.4 million *× μ* = 11, 670 *per* day. Since the incubation period for SARS-CoV-2 (1*/σ*) is estimated to be 3.6 days, we set the transition parameter (*σ*) from the exposed class to the symptomatic or asymptomatic infectious class to be *σ* = 1*/*3.6 *per* day [44]. Using a meta-analysis, Ma *et al*. [45] estimated that 40% of individuals who get infected with COVID-19 become asymptomatically-infectious (this estimate is also consistent with those reported in [46, 47]). Consequently, we set *r* = 0.4. Ferguson *et al*. [48] estimated the duration of recovery for symptomatically-infectious individuals (1*/γ*_*i*_) to be 5 days (hence, *γ*_*I*_ = 1*/*5 *per* day). Niu *et al*. [49] estimated the infectious duration for asymptomatic recovery (1*/γ*_*a*_) to be five days (hence, *γ*_*A*_ = 1*/*5 *per* day). During the time period considered in the study, the highly-effective N95 respirator (with an associated efficacy of *ε*_*m*_ = 0.95) or equivalent [39, 50, 51] were not readily available to the general public (they were prioritized for frontline healthcare workers), we assume that the overall average efficacy of the mask used in the community (i.e., by individuals in group 2) is *ε*_*m*_ = 0.5 (this figure is estimated by averaging the relative efficacy of the surgical (0.7) and cloth (0.3) masks, which were the predominant masks used during the first wave) [31, 39]. Finally, survey data collected for the “COVID States Project” [52] shows the maximum proportion of respondents claiming to wear masks during the survey period of April 2020 to May 2022 was around 80%, hence we estimate that the maximum proportion of new recruitment that adhere to interventions to be 80% (thus, 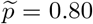). For simplicity, we assumed that mask fatigue was always in play in the United States, *albeit* the level of fatigue increases with the length or duration of the pandemic (thus, we set *t*_*f*_ = 0).

**Table 3:** Values of the fixed parameters of the behavior-epidemiology ((2.4)) and behavior-free ((A.1)) model.

| Parameter | Value | Source |
| --- | --- | --- |
| $\Pi$ | 11,670 day<sup>-1</sup> | [42, 43] |
| $\tilde{p}$ | 0.80 (dimensionless) | [52] |
| $\mu$ | 1/(77.8 × 365) day<sup>-1</sup> | [43] |
| $\varepsilon_m$ | 0.5 (dimensionless) | [31, 39] |
| $r$ | 0.4 (dimensionless) | [45] |
| $\sigma$ | 1/3.6 day<sup>-1</sup> | [44] |
| $\gamma_i$ | 1/5 day<sup>-1</sup> | [49] |
| $\gamma_a$ | 1/5 day<sup>-1</sup> | [55] |
| $t_f$ | 0 (day) | Assumed |
| $\zeta$ | 1/1500 (human<sup>-1</sup>) | Assumed |
| $K$ | 5,000,000 (dimensionless) | Assumed |

#### 2.2.2 Estimated (fitted) parameters of the behavior-epidemiology model

In this section, the values of the unknown parameters of the model will be generated by fitting the model (2.4) with the observed cumulative disease-induced mortality for the SARS-CoV-2 pandemic for the aforementioned period from March 1, 2020 to May 29, 2020 (i.e., during the first wave of the pandemic) in the United States (recall that we set *δ* = *δ*_1_ = *δ*_2_). Nonlinear least-squares optimization method was used to fit the model. MATLAB’s minimization function “lsqcurvefit” is specifically used to minimize the sum of the squared differences between each data point (obtained from Johns Hopkins University COVID-19 repository [53]), and the value of the corresponding cumulative mortality generated by solving the model (2.4). The results obtained, for the model fitting, is depicted in Figure 2(a) with a blue curve. Furthermore, the values of unknown parameters obtained from the data fitting are tabulated in Table 4. The goodness of the fit in Figure 2(a) is assessed by using the fixed and fitted parameters to predict the daily SARS-CoV-2 mortality for the United States during the fitting period and compare this with the actual observed daily SARS-CoV-2 mortality. The results obtained, depicted in Figure 2(b), show a very good fit. The fitted model was cross-validated by comparing the model projection with the observed data for 20 days after the fitting period (i.e., May 30, 2020, to June 18, 2020). The cross-validation period is illustrated by the segment of the curves to the right of the vertical dashed line in Figure 2 (the model’s projection during this period is shown by the green curve). Here, too, Figure 2 shows that the model fits both the cumulative and daily mortality data quite well during the cross-validation period.

**Table 4:** Values of the fitted (estimated) parameters of the behavior-epidemiology model (2.4).

| Parameter | Estimated (fitted) Value |
| --- | --- |
| $\alpha_{12}^c q_{12}^c$ | $9.337 \times 10^{-3}$ day<sup>-1</sup> |
| $\alpha_{21}^c q_{21}^c$ | $9.223 \times 10^{-3}$ day<sup>-1</sup> |
| $\alpha_{12}^i q_{12}^i$ | 0.110 day<sup>-1</sup> |
| $\alpha_{21}^{ns} q_{21}^{ns}$ | $2.597 \times 10^{-3}$ day<sup>-1</sup> |
| $\alpha_{12}^m q_{12}^m$ | 0.305 day<sup>-1</sup> |
| $\alpha_{21}^f q_{21}^f$ | $5.682 \times 10^{-5}$ day<sup>-1</sup> |
| $\beta$ | 0.661 day<sup>-1</sup> |
| $\eta_a$ | 2.275 (dimensionless) |
| $\delta = \delta_1 = \delta_2$ | $2.505 \times 10^{-4}$ day<sup>-1</sup> |

**Figure 2:**
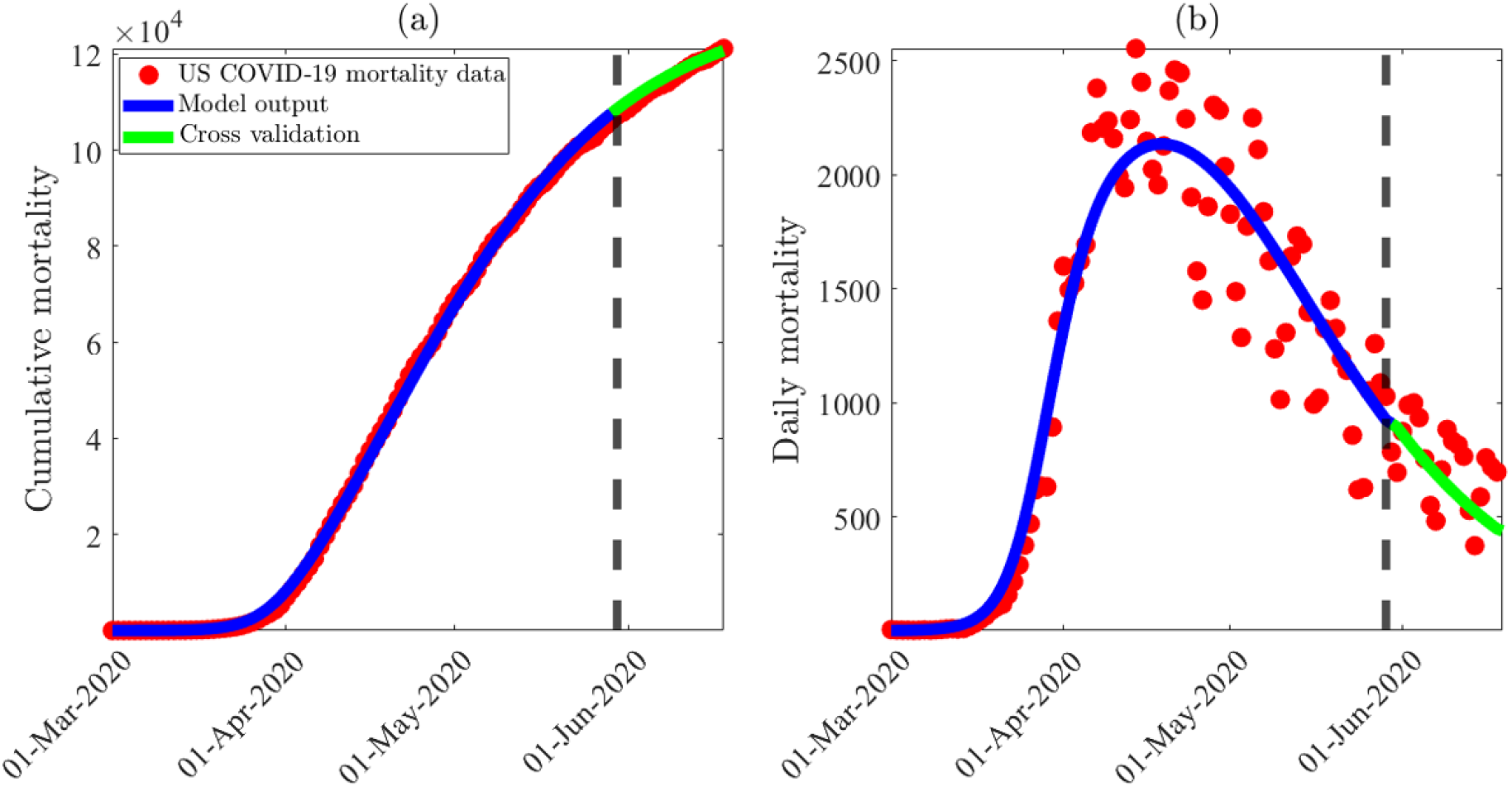
Data fitting, parameter estimation and cross-validation of the behavior-epidemiology model (2.4), using SARS-CoV-2 cumulative mortality data for the United States during the first wave of the pandemic (corresponding to the period from March 1st, 2020 to June 18th, 2020). (a) Least-squares fitting of the behavior-epidemiology model (2.4), showing the cumulative COVID-19 mortality generated from the model, compared to the observed cumulative mortality data for the United States during the first wave (obtained from the Johns Hopkins University COVID-19 repository [53]). (b) Simulation results of the behavior-epidemiology model (2.4), depicting the daily COVID-19 mortality for the United States, as a function of time, predicted by the behavior-epidemiology model using the fixed and estimated parameters tabulated in Tables 3 and 4, respectively, in comparison to the observed daily mortality data for the United States during the first wave (obtained from the Johns Hopkins University COVID-19 repository [53]). The behavior-epidemiology model was fitted to the data from March 1st, 2020 until May 29th, 2020 (depicted by a dashed black line in the figure), and cross-validated by comparing the model output with the observed mortality data for the remaining 20 days (depicted with a green curve). Three of the initial conditions of the behavior-epidemiology model (namely *E*_1_(0), *I*_1_(0) and *A*_1_(0)) were estimated from fitting the model with the aforementioned cumulative mortality data. The estimated values were *E*_1_(0) = 5, 754, *I*_1_(0) = 2, 338 and *A*_1_(0) = 2, 501, respectively. The remaining initial conditions were set to (using US census data near the disease-free equilibrium): *N* (0) = 3.314 *×* 10^8^, *S*_2_(0) = 1, *E*_2_(0) = 0, *I*_2_(0) = 0, *A*_2_(0) = 0, *R*_1_(0) = 0, *R*_2_(0) = 0, *D*(0) = 1, and *S*_1_(0) = *N* (0) − *E*_1_(0) − *I*_1_(0) − *A*_1_(0) − *S*_2_(0).

#### 2.2.3 Initial conditions of the behavior-epidemiology model

The numerical values of the initial conditions used in the simulations of the behavior-epidemiology model (2.4) are described below. First of all, the initial values of some of the state variables of the model for individuals in group 1, notably the number of exposed individuals in group 1 (*E*_1_), the number of symptomatic individuals in group 1 (*I*_1_) and the number of asymptomatic infectious individuals in group 1 (*A*_1_), were obtained from fitting the model with the cumulative mortality data. Specifically, the data fitting shows that the estimated values of *E*_1_(0), *I*_1_(0) and *A*_1_(0) are 5,754, 2,338 and 2,501, respectively. The remaining initial values of the state variables of the model were either obtained from demographic/census data or assumed, as described below. Since the simulations are started at the beginning of the epidemic (i.e., near the disease-free equilibrium), we set the initial cumulative mortality, *D*(0), to *D*(0) = 1 [53], while the initial total population size of the United States is set at *N* (0) = 3.314 *×* 10^8^ [42]. Further, we set the initial number of recovered individuals to zero (i.e., *R*_1_(0) = *R*_2_(0) = 0) and the initial size of the susceptible pool in group 2 is set to 1 (i.e., *S*_2_(0) = 1), while the initial values of the remaining state variables of the model for individuals in group 2 are set to zero (i.e., *E*_2_(0) = *I*_2_(0) = *A*_2_(0) = 0). The initial susceptible population in group 1 (*S*_1_(0)) can be obtained by taking the difference between the initial total population (*N* (0)) and the sum of the initial values of the remaining state variables of the model with a non-zero initial value (i.e., we set *S*_1_(0) = *N* (0) − (*E*_1_(0) + *I*_1_(0) + *A*_1_(0) + *S*_2_(0))).

#### 2.2.4 Insight from fitting the behavior-epidemiology model

The numerical values or sizes of the estimated parameters of the behavior-epidemiology model (2.4), tabulated in Table 4, can provide important insight into the impact of human behavior changes on the trajectory and burden of the disease during the simulation/fitting period (i.e., during the first wave of the pandemic in the United States). In particular, since the estimated value of the modification parameter accounting for the variability in disease transmission by asymptomatically-infectious individuals, in comparison to symptomatically-infectious individuals, is *η*_*a*_ = 2.275, it follows that the transmission rate for asymptomatic infectious individuals (*βη*_*a*_) is at least twice the size of the transmission rate for symptomatic infected individuals (*β*). Furthermore, asymptomatic infectious individuals accounted for at least 70% of all new SARS-CoV-2 cases in the United States during the first wave of the pandemic (this is computed from the ratio: *η*_*a*_ *β/*(*η*_*a*_ *β* + *β*) × 100%). Thus, it follows from the parameter estimation associated with fitting the behavior-epidemiology model with the cumulative SARS-CoV-2 mortality data, that this study confirms that asymptomatic infectious individuals were the main drivers of the SARS-CoV-2 pandemic in the United States during the first wave [29–31]. Table 4 also shows that the estimated value for the product of the maximum rate of behavior change attributed to fatigue to adherence to the public health interventions implemented in the United States during the first wave 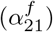, and its associated probability of influence 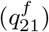, given by 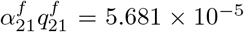 day, is exceptionally small. Hence, it can be inferred (from the very small value of the product 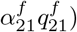 that behavior change due to fatigue to adherence to interventions was negligible (if at all) during the first wave of the pandemic in the United States. Similarly, the estimated values for the product of parameters associated with behavior change attributed to contact (i.e., 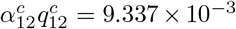 *per* day and 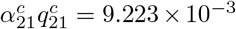*per* day), as well as those for the parameters associated with the proportion of non-symptomatic individuals in the community (i.e., 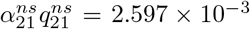 *per* day) were relatively small, suggesting behavior changes due to these metrics (contacts and size of non-symptomatic pools) were of marginal impact on the trajectory or burden of the pandemic during the first wave in the United States.

On the other hand, Table 4 shows that the value of the product of the parameters associated with behavior change due to the level of symptomatic transmission in the community (i.e., 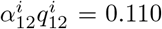 *per* day) and disease-induced mortality (i.e., 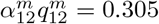 *per* day) were relatively large (in comparison to the sizes of the other estimated parameters). Hence, behavior change due to these metrics (level of symptomatic transmission and disease-induced mortality) play significant role in driving the pandemic during the first wave in the United States (i.e., the most influential forms of behavior change were linked to mortality followed by the level of symptomatic transmission during the first wave of the SARS-CoV-2 pandemic in the United States). The behavior change functions will be further analysed in Section 3.1 (and their impact on disease transmission and disease-induced mortality will also be discussed there).

Finally, it is intuitive to determine how well the model does in capturing the trajectory and burden of the pandemic. This is explored below. It should be, first of all, be recalled that the behavior-epidemiology model was fitted with the cumulative mortality data for the period from March 1, 2020 to May 29, 2020 (shown by the curves to the left of the dashed vertical line in Figure 2). The fitted model was then used to cross-validate the data for the next 20 days, from May 30, 2020 to June 18, 2020 (as shown by the green curve, depicted to the right of the dashed vertical line). Data from the Johns Hopkins University COVID-19 repository [53] (see also Figure 2) shows that the observed cumulative mortality for the SARS-CoV-2 pandemic in the United States at the end of the cross-validation period (i.e., the cumulative mortality as of June 18, 2020) was 121, 131, and the value predicted by the model (green curve for June 18, 2020) is 119, 264 (representing about 1.54% underestimate of the actual data). It should also be stated that during the fitting period (from March 1, 2020 to May 29, 2020), the observed cumulative mortality was 106,290. Since there were 121,132 cumulative deaths at the end of the cross-validation period, it follows that there were 121, 132 − 106, 290 = 14, 842 additional deaths during the cross-validation period. Figure 2 predicts an additional 119, 264 − 106, 290 = 12, 974 deaths during the cross-validation period (which underestimates the corresponding additional mortality by about 12.59%).

The goodness of the fitting depicted in Figure 2 can also be measured in terms of the sum of the squares of the associated residuals, as follows. Let 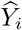 be the observed data (for example, the cumulative mortality or daily mortality) on day *i*, and let *Y*_*i*_ be the model’s output data on day *i*. Thus, the residual on day *i* is 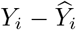 and the sum of squared residuals (SSR), also defined as the sum of squared error (SSE), is given by [54]:

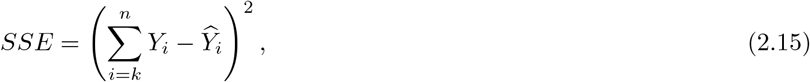

where *k* is the starting point of the sum and *n* is the total number of the given or available (observed) data points. The SSE on the cumulative mortality during the fitting period (i.e., March 1, 2020, to May 29, 2020) and during the cross-validation period (i.e., May 30, 2020, to June 18, 2020) for the model (2.4) was 4.99 *×* 10^7^ and 1.49 *×* 10^7^, respectively. Similarly, the SSE values for the predicted daily mortality during the fitting period (i.e., the period from March 1, 2020, to May 29, 2020) and during the cross-validation period (i.e., the period from May 30, 2020, to June 18, 2020) for the behavior-epidemiology model (2.4) were computed to be 4.87 *×* 10^6^ and 9.02 *×*10^5^, respectively. These values of the SSE will be compared with the corresponding SSE values for the behavior-free equivalent of the model (2.4) (given in Appendix A) to determine which of the two models best fits (i.e., has lower SSE values) the observed cumulative mortality data. That is, the goodness of fit (to the cumulative mortality data in Figure 2(a), and the predicted daily mortality data in Figure 2(b)) will be compared with the goodness of fit for the version of the behavior-epidemiology model (2.4) that does not explicitly incorporate human behavior (given by Equation (A.1) in Appendix A, with the corresponding fits depicted in Figures 3(a) and 3(b), respectively).

**Figure 3:**
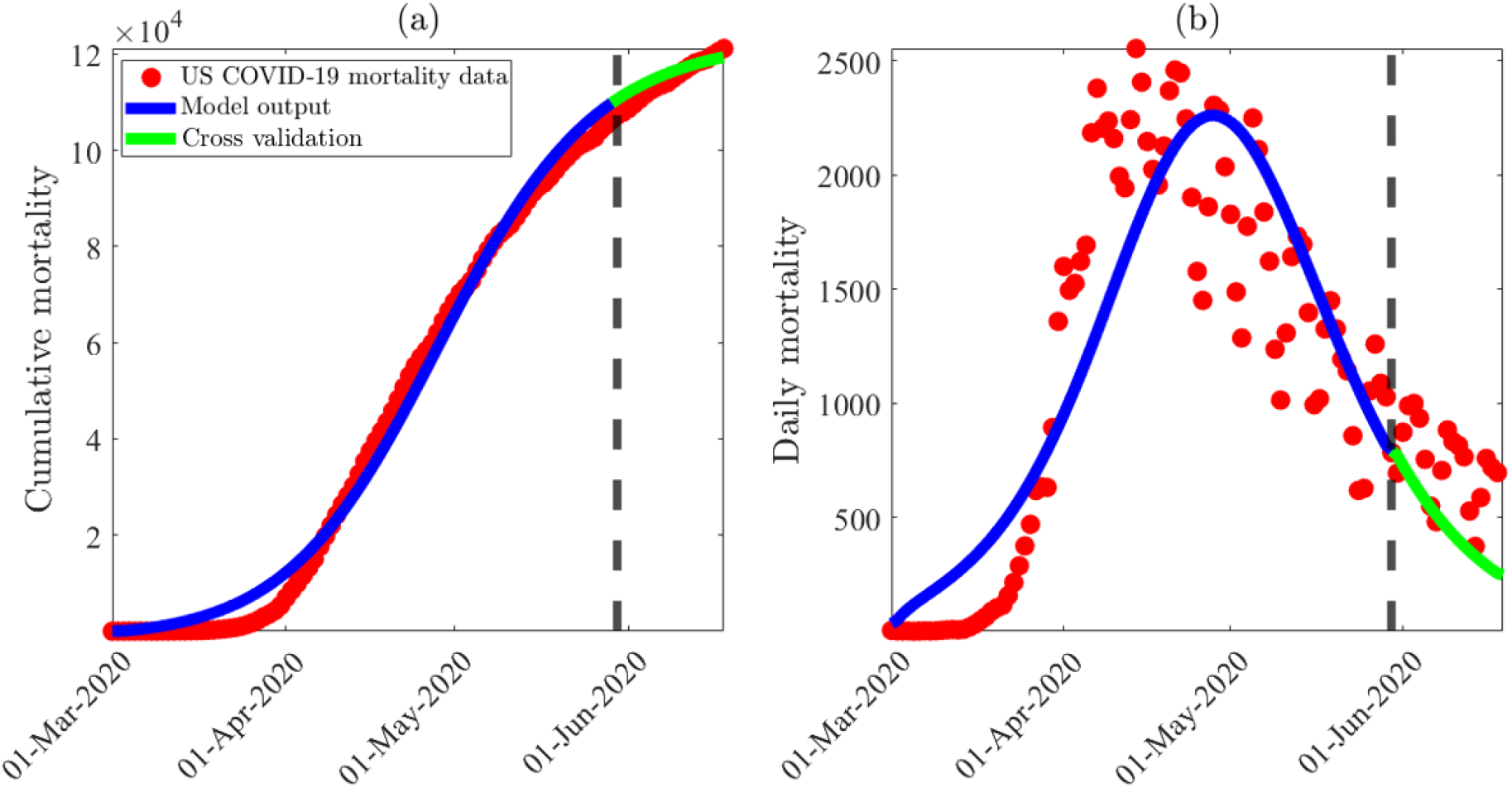
Data fitting, parameter estimation and cross-validation of the behavior-free model (A.1), using SARS-CoV-2 cumulative mortality data for the United States during the first wave of the pandemic (corresponding to the period from March 1st, 2020 to June 18th, 2020). (a) Least-squares fitting of the behavior-free model (A.1), showing the cumulative COVID-19 mortality generated from the model, compared to the observed cumulative mortality data for the United States during the first wave (obtained from the Johns Hopkins University COVID-19 repository [53]). (b) Simulation results of the behavior-free model (A.1), depicting the daily COVID-19 mortality for the United States, as a function of time, predicted by the behavior-free model using the fixed parameters tabulated in Table 3 and estimated parameters obtained through fitting (i.e., *β* = 0.132 *per* day, *η*_*a*_ = 5.349 and *δ* = 1.611 *×* 10^−4^ *per day*), in comparison to the observed daily mortality data for the United States during the first wave (obtained from the Johns Hopkins University COVID-19 repository [53]). The behavior-free model was fitted to the data from March 1st, 2020 until May 29th, 2020 (depicted by a dashed black line in the figure), and cross-validated by comparing the model output with the observed mortality data for the remaining 20 days (depicted with a green curve). Three of the initial conditions of the behavior-free model (namely *E*_1_(0), *I*_1_(0) and *A*_1_(0)) were estimated from fitting the model with the aforementioned cumulative mortality data. The estimated values were *E*_1_(0) = 1, 388, 568, *I*_1_(0) = 10, 3571 and *A*_1_(0) = 350, 357, respectively. The remaining initial conditions were set to (using US census data near the disease-free equilibrium): *S*_2_(0) = 1, *E*_2_(0) = 0, *I*_2_(0) = 0, *A*_2_(0) = 0, *R*_1_(0) = 0, *R*_2_(0) = 0, *D*(0) = 1, *N* (0) = 3.314 *×* 10^8^, and *S*_1_(0) = *N* (0) − *E*_1_(0) − *I*_1_(0) − *A*_1_(0) − *R*_1_(0), *R*_1_(0) = 0.

### 2.3 Fitting the behavior-free model with data

In order to fully understand the impact of explicitly accounting for human behavior elements in the model, it is instructive to also consider the version of the behavior-epidemiology model (2.4) without the behavior elements (known as the *behavior-free model*. The equations for the behavior-free model, obtained by setting all the behavior-related parameters of the behavior-epidemiology model (2.4) to zero (i.e., we consider the model (2.4) with 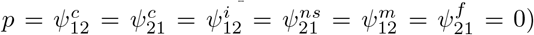 are given by Equation (A.1) in Appendix A. The resulting behavior-free model (A.1) will also be fitted using the same cumulative mortality data used to fit the behavior-epidemiology model and fixed parameters given in Table 3. Here, too, some parameters of the behavior-free model (namely, *β, η*_*a*_ and *δ*), as well as initial conditions (namely *E*_1_(0), *I*_1_(0) and *A*_1_(0)) will be estimated from the data fitting. The remaining initial conditions used in the fitting were (i.e., the same as used for the corresponding state variables of the behavior-epidemiology model): *N* (0) = 3.314 *×* 10^8^, *S*_2_(0) = 1, *E*_2_(0) = 0, *I*_2_(0) = 0, *A*_2_(0) = 0, *R*_1_(0) = 0, *R*_2_(0) = 0, *D*(0) = 1, and *S*_1_(0) = *N* (0) − *E*_1_(0) − *I*_1_(0) − *A*_1_(0) − *S*_2_(0). The results obtained, for fitting the behavior-free model, are depicted in Figure 3, and the values of the estimated parameters (obtained from fitting the behavior-free model) were obtained to be *β* = 0.152 *per* day, *η*_*a*_ = 5.349 and *δ* = 1.611 *×* 10^−4^ *per day*. Similarly, the estimated values of the initial conditions were: *E*_1_(0) = 1, 388, 568, *I*_1_(0) = 10, 3571 and *A*_1_(0) = 350, 357, respectively. Figure 3 also shows a general qualitative trend, but with some important differences, in comparison to the results of the fitting obtained for the behavior-epidemiology model (2.4), as described below.

#### 2.3.1 Insights from fitting the behavior-free model (A.1) with data

First of all, since the estimated value of the modification parameter accounting for the variability in disease transmission by asymptomatically-infectious individuals (*η*_*a*_), for the behavior-free model, is *η*_*a*_ = 5.41, it follows that, for the behavior-free model (A.1), the transmission rate for asymptomatic infectious individuals (*βη*_*a*_) is at least five times as much as that of symptomatic infected individuals (*β*). Thus, the behavior-free model shows that asymptomatic infectious individuals are a lot more influential (in generating more new cases), in comparison to the model with human behavior (in other words, adding heterogeneity due to human behavior changes during an epidemic significantly reduces the relative infectiousness or influence of asymptomatic transmission). Furthermore, specifically for this model, the fitting shows that asymptomatic infectious individuals accounted for about 84% (*η*_*a*_*β/*(*η*_*a*_*β* + *β*) *×* 100%) of all new cases during the first wave of the pandemic in the United States (recall, from Subsection 2.2.2, that asymptomatic individuals accounted for 70% of new cases using the behavior-epidemiology model (2.4)). Since most of the modeling and empirical studies suggest that asymptomatic individuals accounted for between 50% to 70% of new cases [56–58], this study shows that the model without human behavior (unlike with human behavior) over-estimated the influence of asymptotic transmission in the United States.

It should be recalled that the model was fitted with data for the period from March 1, 2020 to May 29, 2020 (shown by the curves to the left of the dashed vertical line in Figure 3). The fitted model was then used to cross-validate the data for the next 20 days, May 30, 2020 to June 18, 2020 (as shown by the green curve, depicted to the right of the dashed vertical line). Data from the Johns Hopkins repository [53] (see also Figure 3) showed that the observed cumulative mortality for the SARS-CoV-2 pandemic in the United States at the end of the cross-validation period (i.e., the cumulative mortality as of June 18, 2020) was 121,131, and the value predicted by the model (green curve for June 18, 2020) is 115,865 (representing about 4.35% underestimate of the actual data). Recall that the behavior-epidemiology model (2.4) underestimated this data by about 1.54%. It should also be stated that during the fitting period (from March 1, 2020 to May 29, 2020), the observed cumulative mortality was 106,290. Since there were 121,132 cumulative deaths at the end of the cross-validation period, it follows that there were 121, 132 − 106, 290 = 14, 842 additional deaths during the cross-validation period. Figure 3 predicts an additional 115, 866 − 106, 290 = 9, 576 deaths during the cross-validation period, which represents an underestimation of the observed additional mortality by about 35.49%. It is worth recalling, from Section 2.2.4, that the behavior-epidemiology model (2.4) only underestimated this data (the additional mortality during the cross-validation period) by about 12.59%. In other words, the behavior-epidemiology did far better in predicting the additional mortality during the cross-validation period than the corresponding behavior-free model.

Furthermore, the SSE computed from fitting the behavior-free model (A.1) (computed using Equation (2.15)) with the cumulative mortality during the fitting period (i.e., March 1, 2020, to May 29, 2020) and the cross-validation period (i.e., May 30, 2020, to June 18, 2020), depicted in Figure 3(a), was 7.81 *×* 10^8^ and 1.36 *×* 10^8^, respectively. These SSE values are 15.65 and 9.13 times larger than the corresponding SSE values for the behavior-epidemiology model (given in Section 2.2.4). Similarly, the SSE value computed for the daily mortality predicted by the behavior-free model during the fitting and cross-validation periods, depicted in Figure 3(b), are 1.31 10^7^ and 3.12 10^6^, respectively (these SSE values are 2.69 and 3.46 times larger than those obtained for the behavior-epidemiology model (2.4), given in Section 2.2.4)). Hence, in summary, these goodness of fit computations show that the behavior-epidemiology model (2.4) does far better in fitting the observed cumulative mortality data during the fitting and cross-validation periods, in addition to providing a more accurate prediction of the daily mortality observed during these period. It is worth noting (from Sections 2.2.4 and 2.3.1), however, that the SSE values for the cumulative mortality are always larger than those for the daily mortality predicted by the two models (this is to be expected, considering the “filing up” effect of cumulative numbers, in relation to daily numbers [59]. Furthermore, as stated earlier, the SSE values for the behavior-epidemiology model (2.4) (for both cumulative and daily mortality mortality) is always lower than the corresponding ones generated using the behavior-free model, emphasizing the superiority of the behavior-epidemiology model to better capture the observed trajectory (and burden) of the disease and predicts its future trajectory (and burden). The SSE values, together with the other statistical metrics for measuring the goodness of fit for the two models (notably the root mean squared error (RMSE) and root mean squared logarithmic error (RMSLE)) are tabulated in Table B.1 and B.2 of Appendix B. This table shows that the behavior-epidemiology model consistently has lower errors than the behavior-free model.

Figure B.1 show the correlation between the reported daily mortality and the predicted daily mortality generated using the behavior-epidemiology (Figure B.1(a)) and the behavior-free models (Figure B.1(b)), from which it can be seen that the correlation coefficients (*R*) and the r-squared (*R*^2^) [60] associated with the behavior-epidemiology model (*R* = 0.96 and *R*^2^ = 0.92) are closer to one than those for the behavior-free model (*R* = 0.88 and *R*^2^ = 0.77), signifying that the behavior-epidemiology model more accurately captures the daily mortality trend. Hence, it can be concluded, based on the visual goodness of fit, the difference in projected mortality, and the aforementioned statistical metrics, that the behavior-epidemiology model (2.4) performed better in capturing the trend and projecting the trajectory of both cumulative and daily mortality during the first wave of COVID-19 in the United States compared to the behavior-free model (A.1).

### 2.4 Basic qualitative properties of the behavior-epidemiology model

Before analysing the asymptotic properties of the behavior-epidemiology model (2.4), it is instructive to analyse its basic qualitative properties first of all. This is done below. We define the following biologically-feasible region for the model:

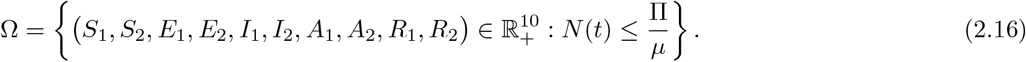

#### Theorem 2.1.

*Suppose that the initial values S*_1_(0), *S*_2_(0), *E*_1_(0), *E*_2_(0), *I*_1_(0), *I*_2_(0), *A*_1_(0), *A*_2_(0), *R*_1_(0), *R*_2_(0) *of the model* (2.4) *are non-negative. The region* Ω *is positively-invariant and bounded with respect to the model* (2.4).

*Proof*. The equation for the rate of change of the total human population, obtained by adding all equations in (2.4), is given by:

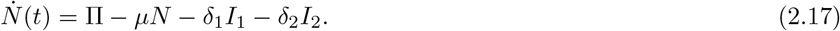

By the non-negativity of the parameters and state variables of the model (2.4), it follows from (2.17) that

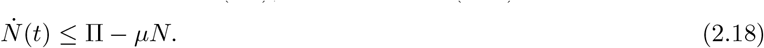

Hence, if 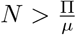, then 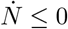. Thus, it can be shown using a standard comparison theorem [61] on (2.18) that

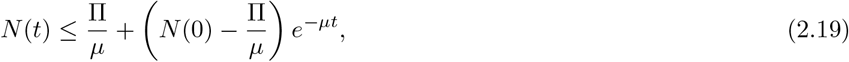

where *N* (0) represents the initial population (i.e., the population at time *t* = 0). Hence, 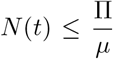 if 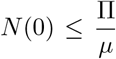. If the initial population exceeds the carrying capacity 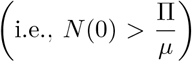 then the solutions are initially outside the region Ω but the solution trajectory will eventually enter the region Ω since 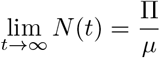. Thus, every solution of the model (2.4) with initial conditions in Ω remains in Ω for all time *t >* 0, and those outside Ω are eventually attracted into Ω. In other words, the region Ω is positively-invariant and attracts all initial solutions of the model (2.4).

### 2.5 Asymptotic stability of disease-free equilibria

The model (2.4) has two disease-free equilibria (DFE), namely a trivial *adherents-free* DFE (denoted by 𝔼_1_) and a non-trivial *adherents-present* DFE (denoted by 𝔼_2_), given, respectively, by:

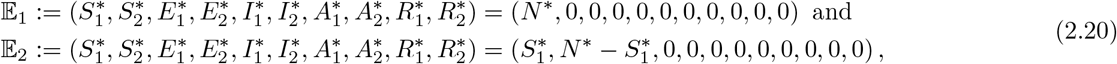

where (for 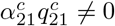 and *r*_0_*≠*1),

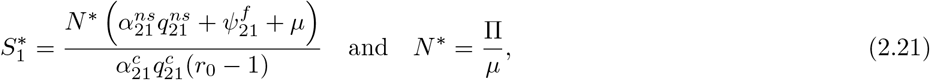

with 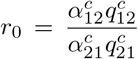 (for 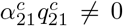). It follows from (2.21) that the DFE E_2_ exists if and only if *r*_0_ *>* 1. This result is summarized below.

#### Theorem 2.2.

*The behavior-epidemiology model* (2.4) *has a trivial disease-free equilibrium (𝔼*_1_, *which always exists) and a non-trivial disease-free equilibrium (𝔼*_2_*), which exists whenever r*_0_ *>* 1.

The quantity *r*_0_ is the ratio of the *contact-mediated* capacity of non-adherent individuals to become adherents (at the rate 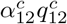) to that of adherent individuals becoming non-adherents (at the rate 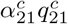). In other words, *r*_0_ *>* 1 implies that non-adherent individuals adhere to interventions and move to the adherents group at a rate faster than that at which adherent individuals become non-adherent and move to the non-adherent group (i.e., *r*_0_ *>* 1 means the overall rate of transition from group 1 to group 2 exceeds that for group 2 to group 1). The trivial disease-free equilibrium (𝔼_1_) is not epidemiologically realistic, since it does not have any individuals in group 2 (hence, it will not be considered or analysed in this study).

#### 2.5.1 Local asymptotic stability of the non-trivial DFE of the behavior-epidemiology model

In this section, the *next generation operator method* [62, 63] will be used to analyse the local asymptotic stability property of the non-trivial disease-free equilibrium (𝔼_2_) of the behavior-epidemiology model (2.4). The implementation of the next generation operator method requires the computation of two matrices, one that keeps track of new infection terms (denoted by *F*) and the other that tracks the linear transition terms (denoted by *V*). It can be shown, using the notation in [62], that the matrices *F* and *V*, corresponding to the behavior-epidemiology model (2.4), are given, respectively, by:

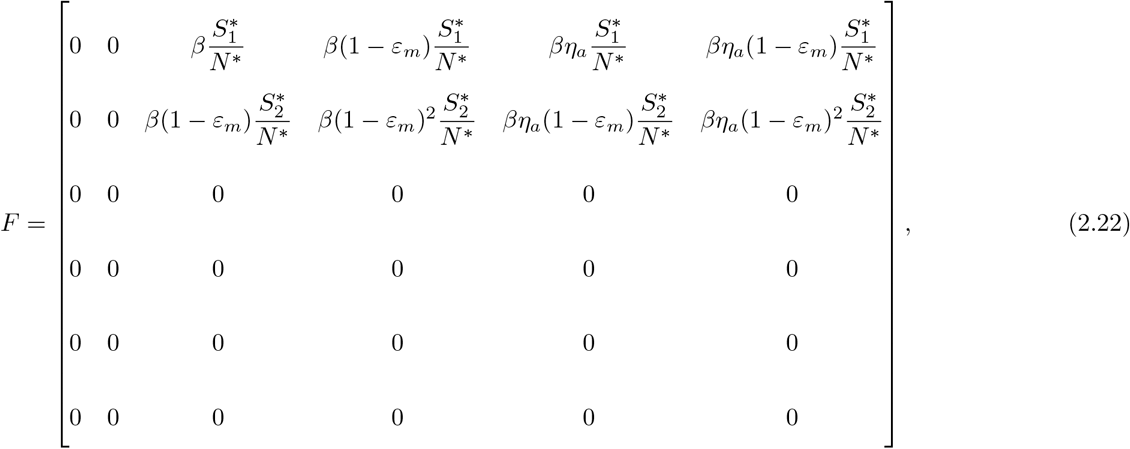

and,

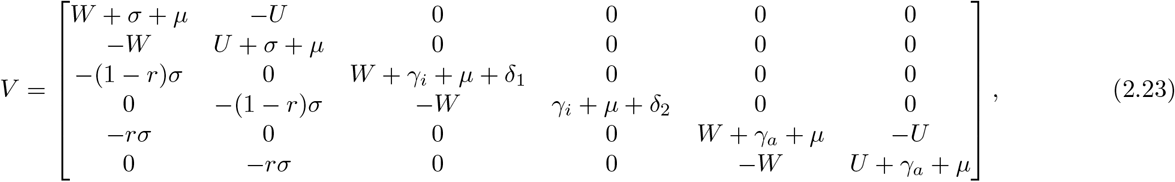

where,

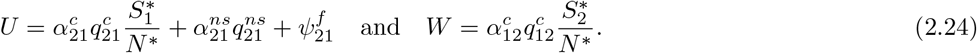

It is convenient to define the following quantity (where *ρ* is the spectral radius):

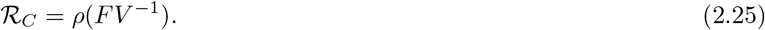

The result below follows from Theorem 2 of [62] (note that closed form expression for ℛ_*C*_ is not readily available, due to the high dimensionality of the associated next generation matrices, and has to be computed numerically).

##### Theorem 2.3.

*The non-trivial disease-free equilibrium of the behavior-epidemiology model* (2.4) *(which exists only if r*_0_ *>* 1*) is locally-asymptotically stable if ℛ*_*C*_ *<* 1, *and unstable whenever ℛ*_*C*_ *>* 1.

The quantity ℛ_*C*_ is the *control reproduction number* of the behavior-epidemiology model (2.4). It measures the average number of new cases generated by a typical infectious individuals if introduced in a population where some public health intervention measures (notably nonpharmaceutical interventions, such as the use of face mask in public) are implemented. The epidemiological implication of Theorem 2.3 is that a small influx of infected individuals into the community will not generate a large outbreak in the community if the control interventions can bring (and maintain) the threshold quantity ℛ_*C*_ to a value less than one. Using the baseline values of the fixed and estimated parameters in Tables 3 and 4 in Equation (2.25) shows that the value of ℛ_*C*_ during the first wave of the SARS-CoV-2 pandemic in the United States was ℛ_*C*_ ≈ 2.42 (showing that the non-trivial equilibrium is unstable, and suggesting significant outbreak of the disease). It is worth stating that the value of the control reproduction number associated with the behavior-free model (A.1), denoted by 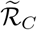 and computed by substituting the parameters in Table 3 and the estimated parameters for the behavior-free model (namely *β* = 0.132 *per* day, *η*_*a*_ = 5.349 and *δ* = 1.611 *×* 10^−4^ *per* day) into the corresponding expression for its associated control reproduction number, is 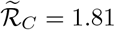. Most modeling studies have suggested a control reproduction number for the first wave of SARS-CoV-2 in the United States in the range [2-4] [64, 65]. Thus, the behavior-free model, unlike the behavior-epidemiology model, may have under-estimated the burden (or severity) of the first wave in the United States.

## 3 Numerical Simulations of Behavior-Epidemiology Model

The behavior-epidemiology model (2.4) will now be simulated, using the baseline values of the fixed and estimated parameters of the model tabulated in Tables 3 and 4, to assess the impacts of disease-related metrics (namely, contacts with individuals from another group, level of disease-induced mortality, level of symptomatic transmission, proportion of non-symptomatic individuals and intervention fatigue in the community) on inducing behavior change (as measured in terms of the back-and-forth transitions between the two behavior groups considered in this study) during the first wave of the SARS-CoV-2 pandemic in the United States. Simulations will also be carried to assess the impact of the various behavior change functions and changes in the value of parameter that accounts for the proportion of exposed individuals who become asymptomatically-infectious at the end of the exposed period (*r*) on disease burden (specifically, cumulative mortality and cumulative) during the first wave. It should be emphasized that the aforementioned proportion *r* (of asymptomatic individuals) can change over time due to numerous factors, such as increases in population-wide immunity due to reinfection or the emergence of a new variant (a systematic review by Yu *et al*. [66] showed that the proportion of individuals infected with the Omicron variant that were asymptomatically-infectious was much higher, in comparison to those infected with the Delta variant). Hence, it is crucial to assess (through numerical simulation) the impact of changes in the proportion of exposed individuals who become asymptomatically-infectious at the end of exposed period (*r*) on the disease trajectory, burden and human behavior with respect to adherence, or lack thereof, to public health interventions. These simulations are described below.

### 3.1 Relative influence of disease metrics on inducing behavior change in the community

In this study, behavior change is measured in terms of the transition from the adherent (non-adherent) to the non-adherent (adherent) group. These transitions, or associated behavior change functions (*ψ*_*ij*_; *i, j* = {1, 2 }; *i ≠ j*), are influenced by the following disease metrics: contacts with individuals from another group (captured by the functions 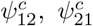 given by Equations (2.8), (2.9), respectively), the level of disease transmission by symptomatic individuals (represented by 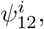 given by (2.10)), the level of non-symptomatic individuals in the community (represented by 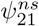, given by (2.11)), the level of disease-induced mortality (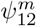, given by (2.12)) and the level of intervention fatigue (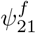, defined in (2.13)) in the community. The behavior change functions 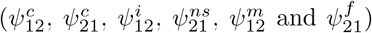 are depicted in Figure 4, from which it follows that behavior change (as measured in terms of transition from group 1 to group 2 or vice versa) is most influenced by two disease metrics: the level of disease-induced mortality in the community (red curve in Figure 4(a)) followed by the level of symptomatic transmission (blue curve in Figure 4(a)). As shown in Figure 4(b)), behavior change from group 2 to group 1 due to contact is more influential during the very early stage of the first wave (brown curve in Figure 4(b)). As the outbreak progresses, however, this mechanism of behavior change loses influence to, for example, behavior change due to contact with individuals in group 2 (light blue curve in Figure 4(b)). Additionally, it is also observed that behavior change due to the presence of non-symptomatic individuals in the community (gold curve in Figure 4(b)) is slightly lower when the number of symptomatic cases significantly increase but otherwise remains nearly constant throughout the first wave). Behavior change due to intervention fatigue was of very marginal (or no) influence during the first wave (dark blue curve in Figure 4(b)). In summary, the simulations in Figure 4 show that the level of disease-induced mortality and disease transmission by symptomatic individuals were the main drivers for positive behavior change from group 1 (non-adherent) to group 2 (adherent) during the first wave of the SARS-CoV-2 pandemic in the United States. The disease metric related to contact and the proportion of non-symptomatic individuals in the community were largely of marginal impact while the metric related to fatigue being inconsequential in inducing significant behavior change during the first wave of COVID-19 in the United States.

**Figure 4:**
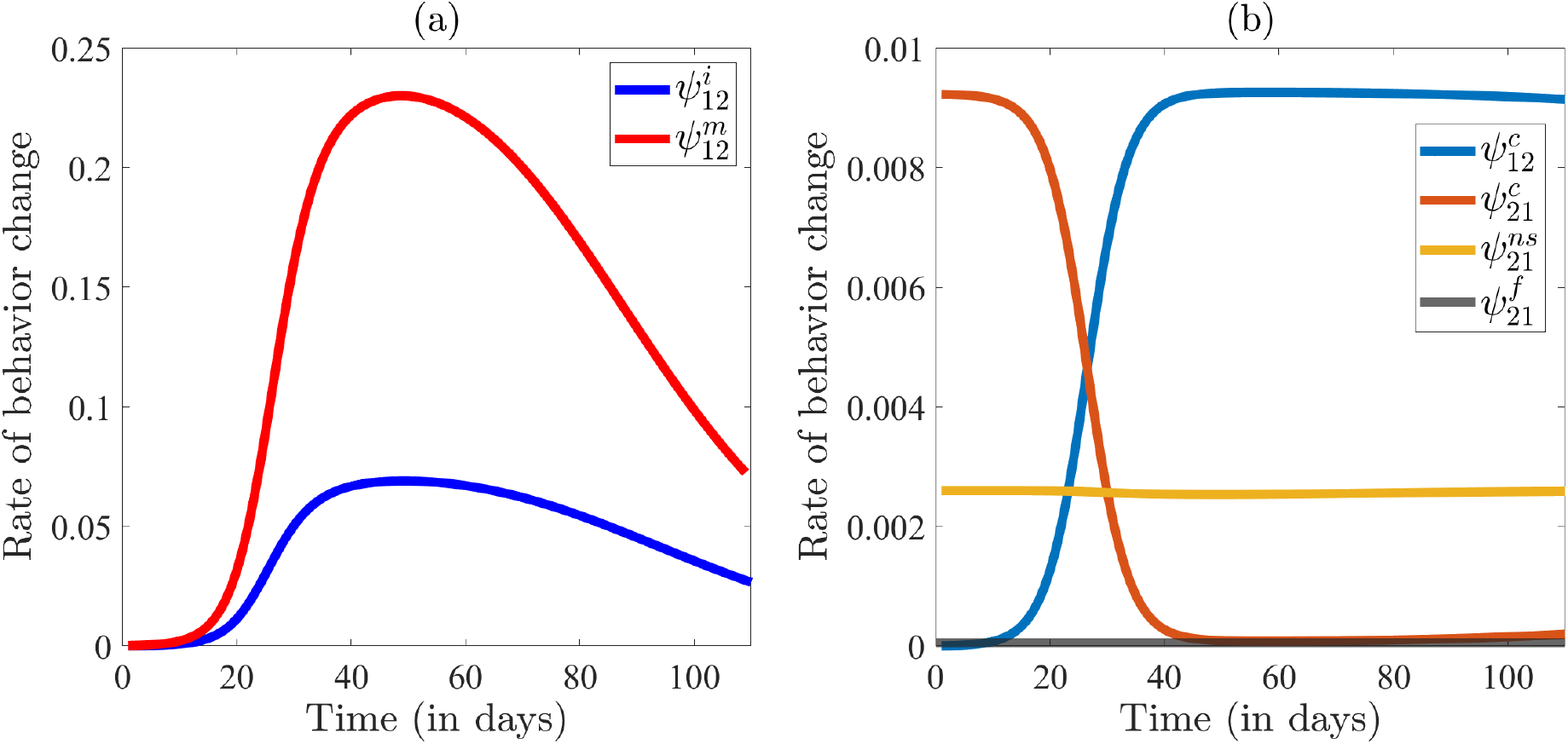
Simulations of the behavior-epidemiology model (2.4) assessing the impact of disease-related metric on behavior change during the first wave of COVID-19 in the United States. The rate of behavior change (a) due to infection 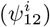 and mortality 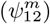 and (b) due to contact with individuals in group 2 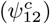, contact with individuals in group 1 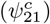, presence of non-symptomatic individuals in the community 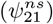 and fatigue 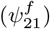 plotted as a function of time. The figure is generated using baseline parameter values from Tables 3 and 4.

### 3.2 Effect of behavior change on mortality

In this section, the behavior-epidemiology model (2.4) is simulated to assess the impact of the five behavior change metrics (namely metrics with respect to contacts with members of the other group, level of mortality, symptomatic transmission, proportion of non-symptomatic individuals and intervention fatigue level in the community) on disease burden (specifically, cumulative mortality). The results obtained for the cumulative mortality are depicted in Figure 5. For these simulations, the product of the respective maximum behavior change rate (*α*_*ij*_) and the associated probability of influence in individuals in group *j* to move to group *i* (*q*_*ij*_) are multiplied by 2-fold (magenta curve), 5-fold (green curve) and 10-fold (red curve) and are compared with the baseline scenario (blue curve).

**Figure 5:**
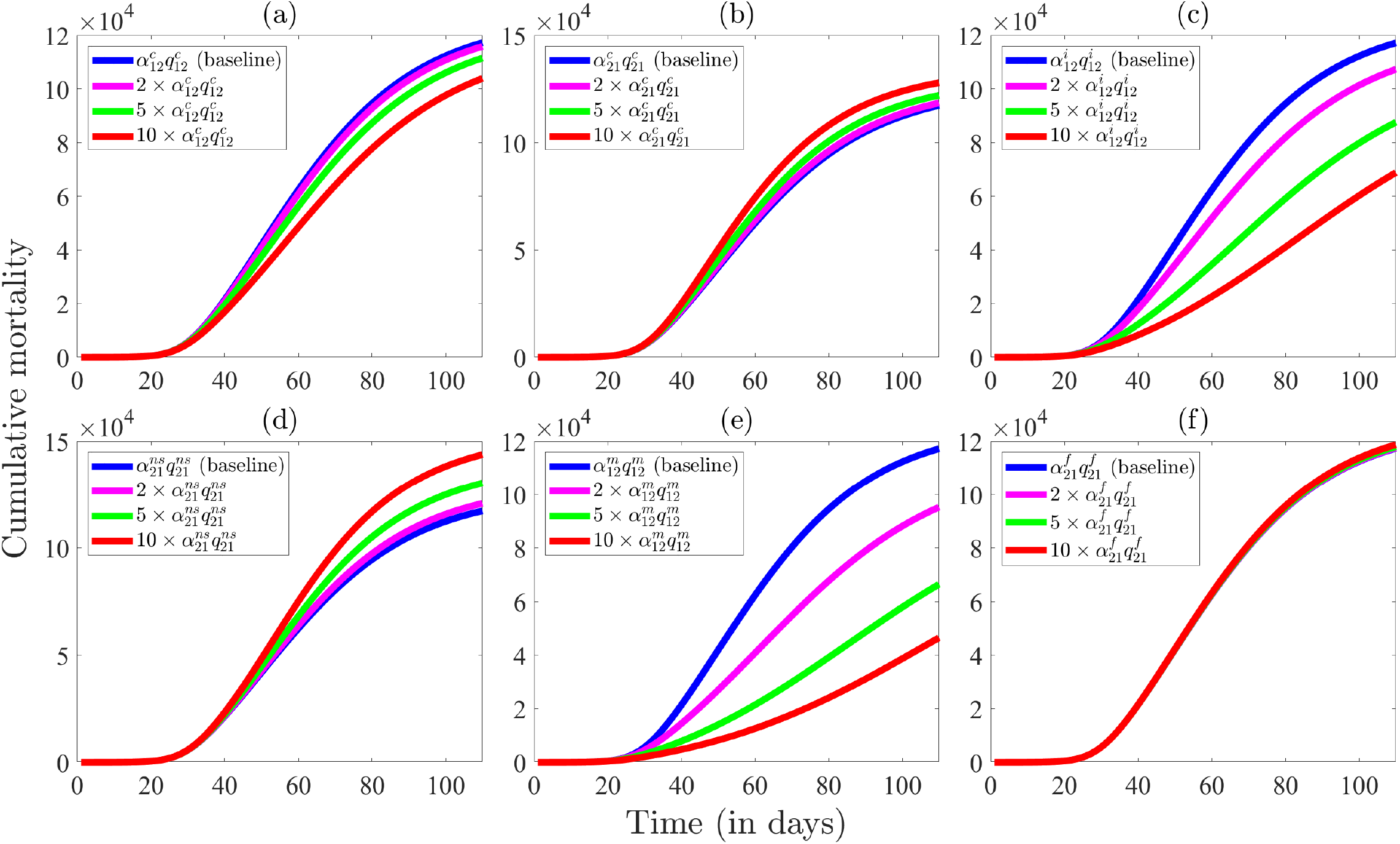
Simulations of the behavior-epidemiology model (2.4) assessing the impact of behavior change metrics on the cumulative mortality as a function of time. The impact on cumulative mortality is shown by increasing the product of the maximum rate of behavior change (*α*_*ij*_) and its associated parameter of influence (*q*_*ij*_) by 2-fold, 5-fold and 10-fold and comparing the obtained cumulative mortality with that of the baseline scenario. The impact on cumulative mortality is assessed by amplifying the above-mentioned parameters related to behavior change due to (a) contact with individuals in group 2 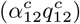, (b) contact with individuals in group 1 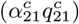, (c) level of symptomatic transmission 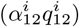, (d) proportion of non-symptomatic individuals in the community 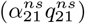, (e) disease-induced mortality 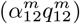 and (f) fatigue to intervention 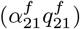 by 2-fold, 5-fold and 10-fold from their baseline values. The baseline figure (i.e., cumulative mortality generated using parameter values from Tables 3 and 4) is depicted with a blue curve while magenta, green, and red curves project cumulative mortality when the respective behavior change parameters are amplified by 2-fold, 5-fold and 10-fold, respectively, with remaining parameter held at their baseline values.

Figure 5 shows that behavior change in favor of transition from the non-adherent group (group 1) to the adherent group (group 2) due to contact individuals in group 1 have with those in group 2 (as measured by 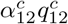) reduces cumulative mortality (albeit marginally) with increasing levels of the overall transition rate 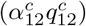, in comparison to the baseline scenario (Figure 5(a)). On the other hand, transition from group 2 to group 1 due to contacts individuals in group 2 have with those in group 1 (as measured by the product 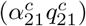 increases cumulative mortality (albeit marginally), in relation to the baseline (Figure 5(b)). Far more significant reductions in cumulative mortality are recorded for the behavior change transitions from group 1 to group 2 due to the level of symptomatic transmission (Figure 5(c)) and disease-induced mortality (Figure 5(e)) in the community, with the latter resulting in far more significant reduction. Furthermore, while the behavior change transition from group 2 to group 1 due to the size of the proportion of non-symptomatic individuals also (marginally) increases cumulative mortality (Figure 5(d)), behavior change due to the level of intervention fatigue has essentially no effect on cumulative mortality in the community during the first wave (Figure 5(f)). These simulations show that the behavior change metric that increases cumulative mortality the most is the change that induces transition from group 2 to group 1 due to the size of non-symptomatic individuals in the community (Figure 5(d)), followed by the behavior change from group 2 to group 1 due to contact (Figure 5(b)). Thus, individuals in group 2 could let their guard down (and transition to group 1) based on the rising levels of non-symptomatic individuals in the community and/or contacts they have with individuals in group 1, the thereby causing more infections and mortality. On the other hand, behavior change from group 1 to group 2 due to mortality has the highest impact in reducing cumulative mortality (Figure 5(e)), followed by corresponding behavior change due to level of symptomatic transmission (Figure 5(c)). In summary, the simulations in Figure 5 show that (positive) behavior change due to level of disease-induced mortality and symptomatic transmission greatly reduces cumulative mortality during the first wave of the SARS-CoV-2 pandemic in the United States, while (negative) behavior change due to level of non-symptomatic individuals or contact with individuals in group 1 increases cumulative mortality during the first wave of the pandemic in the United States.

### 3.3 Impact of size of proportion of exposed individuals who become asymptomatically-infectious at the end of exposed period (*r*)

Asymptomatic infectious individuals were shown, in Section 2.2.4, to account for a sizable proportion of new SARS-CoV-2 cases during the first wave in the United States [29–31]. In this section, the impact of the parameter accounting for the proportion of exposed individuals that become asymptomatic after the exposed period (*r*) on disease burden and inducing behavior change will be assessed.

#### 3.3.1 Impact of the proportion of exposed individuals who become asymptomatically-infectious at the end of exposed period (*r*) on control reproduction number (ℛ_*C*_)

The objective here is to quantify the impact of changes in the proportion of exposed individuals who become asymptomatic at the end of the exposed period (*r*) on disease burden (as measured in terms of the value of the control reproduction number, ℛ_*C*_ of the behavior-epidemiology model (2.4)), (given by Equation (2.25)). Figure 6(a) depicts a profile of ℛ_*C*_, as a function of *r*, generated by using the baseline values of the parameters in Tables 3 and 4, from which it follows that the control reproduction number (hence, disease burden) increases with increasing values of the proportion of exposed individuals who become asymptomatically-infectious at the end of the exposed period (*r*). This is intuitive, since asymptomatic infectious individuals are expected to have more contacts in the community, thereby generating more new cases (in comparison to symptomatic individuals, who may be bed-ridden or in self-isolation; thereby having limited or no contact with the public). However, when the value of the parameter for the relative infectiousness of asymptomatic individuals, in relation to symptomatic infectious individuals (*η*_*a*_) is reduced from its estimated baseline value of *η*_*a*_ = 2.275 to *η*_*a*_ = 0.85, for example, while all other parameters are kept at their baseline values in Tables 3 and 4, the result obtained show that the control reproduction number (ℛ_*C*_) decreases with increasing values of *r* (Figure 6(b)). Hence, this result shows that if the relative infectiousness of asymptomatic infectious individuals is lower than that of symptomatic individuals (for example if asymptomatic individuals have lower contacts or lower viral load and transmission capacity, in comparison to symptomatic individuals), then an increase in the proportion of exposed individuals who are asymptomatic can reduce disease burden. In other words, this study shows that the size of asymptomatic infectious individuals (as measured by the parameter *r*) could lead to an increase or a decrease in disease burden in the community, depending on whether or not the relative infectiousness of asymptomatic infectious individuals is larger or lower than the relative infectiousness of symptomatic infectious individuals (i.e., this depends on whether *r >* 1 or *r <* 1).

**Figure 6:**
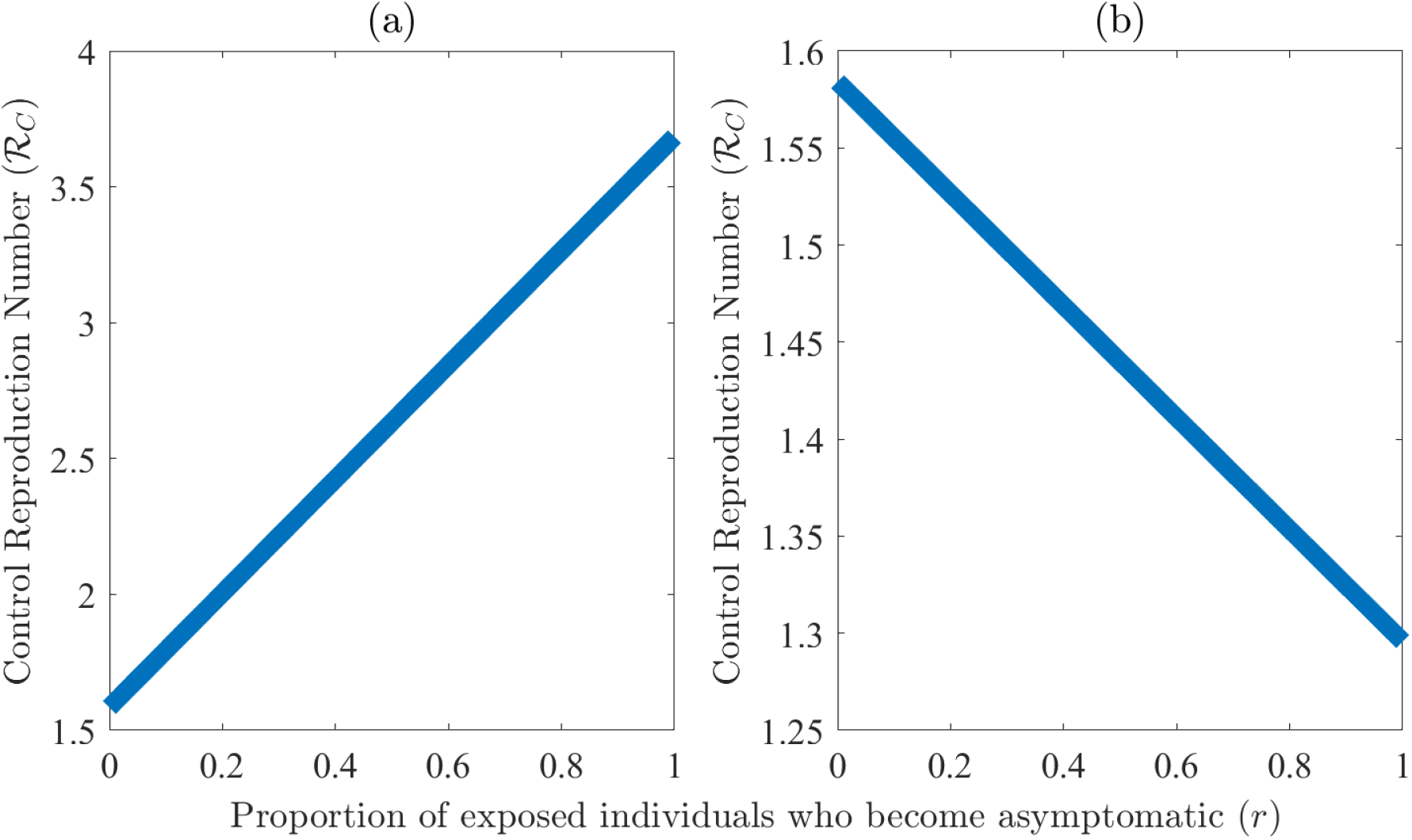
Simulations of the behavior-epidemiology model (2.4) assessing the impact of change of the proportion of exposed individuals who become asymptomatic at the end of the exposed period (*r*) on the control reproduction number (*ℛ*_*C*_). An increase in *r* can (a) increase or (b) decrease *ℛ*_*C*_. Figure (a) is generated by calculating *ℛ*_*C*_, given by Equation (2.25), for varying values of *r* between 0 and 1, with remaining parameter values as in Tables 3 and 4. Figure (b) is similarly generated by varying *r* between 0 and 1, *η*_*a*_ = 0.8 and remaining parameter values as in Tables 3 and 4.

#### 3.3.2 Impact of the proportion of exposed individuals who become asymptomatically-infectious at the end of exposed period (*r*s) on mortality

The objective here is to quantify the impact of changes in the proportion of exposed individuals who become asymptomatic at the end of the exposed period (*r*) on SARS-CoV-2 mortality during the first wave in the United States. Here, too, the behavior-epidemiology model (2.4) is simulated using the baseline parameter values given in Tables 3 and 4, with varying values of *r*. The results obtained are depicted in Figure 7. The profile of the cumulative SARS-CoV-2 mortality in the United States during the first wave, as a function of *r* is depicted in Figure 7(a). This figure show that the cumulative mortality increases with increasing values of *r* until a peak is reached (at about *r* = 0.46), above which the cumulative mortality decreases, reaching zero at *r* = 1 (this is intuitive since *r* = 1 corresponds to the case that all exposed individuals become asymptomatic at the end of the exposed period, and, in the formulation of the behavior-epidemiology model, it was assumed that asymptomatic infectious individuals do not suffer disease-induced mortality). Thus, this study shows the existence of a critical threshold for the proportion of exposed individuals who become asymptomatic at the end of the exposed period that maximizes the cumulative SARS-CoV-2 mortality during the first wave in the United States. Specifically, if *r* = 0.46 (i.e., 46% of exposed individuals do not show clinical symptoms of SARS-CoV-2 at the end of the exposed period, while the remaining 54% do so), cumulative SARS-CoV-2 mortality in the United States is maximized. Above this threshold value, the cumulative SARS-CoV-2 mortality significantly decreases (reaching zero when all exposed individuals do not show symptoms of the disease at the end of the exposed period). This result shows that a strategy that significantly increases the proportion of exposed individuals who become asymptomatic at the end of the exposed period (e.g., rapid testing, detection and treatment of exposed (i.e., newly-infected but not yet infectious) individuals) will significantly reduce the cumulative mortality of SARS-CoV-2 during the first wave in the United States. Furthermore, the behavior-epidemiology model is further simulated using the baseline parameter values in Tables 3 and 4 to generate the profile of the maximum daily SARS-CoV-2 mortality during the first wave of the pandemic in the United States, as a function of the proportion of exposed individuals who become asymptomatic at the end of the exposed period (*r*). The results obtained, depicted in Figure 7(b), also show an increase in the maximum daily mortality with increasing values of *r* until a peak is reached at *r* = 0.64, and the maximum daily mortality decreases thereafter (in other words, simulating the behavior-epidemiology model with *r* = 0.64, and all other parameters as given in Tables 3 and 4, resulted in the highest peak in the daily SARS-CoV-2 mortality during the first wave). Thus, based on the parameter values used in the simulations in this section, the current study identifies two distinct threshold values of the parameter *r* (for the proportion of exposed individuals that become asymptomatic at the end of the exposed period) that maximize cumulative and daily mortality, respectively. While the daily SARS-CoV-2 mortality is maximized during the first wave when the proportion of exposed individuals who become asymptomatic at the end of the exposed period is 64% (this is equivalent to maximizing the mortality burden on the healthcare system if measured on a daily basis), the cumulative mortality is maximized when the proportion is 46% (this is equivalent to maximizing the overall mortality burden on the healthcare burden from the beginning of the pandemic until the end of the first wave). The profiles of the cumulative and daily SARS-CoV-2 mortality, as functions of time, for the two threshold values of *r* (i.e., *r* = 0.46 and *r* = 0.64) are depicted in Figure C.1 (in Appendix C).

**Figure 7:**
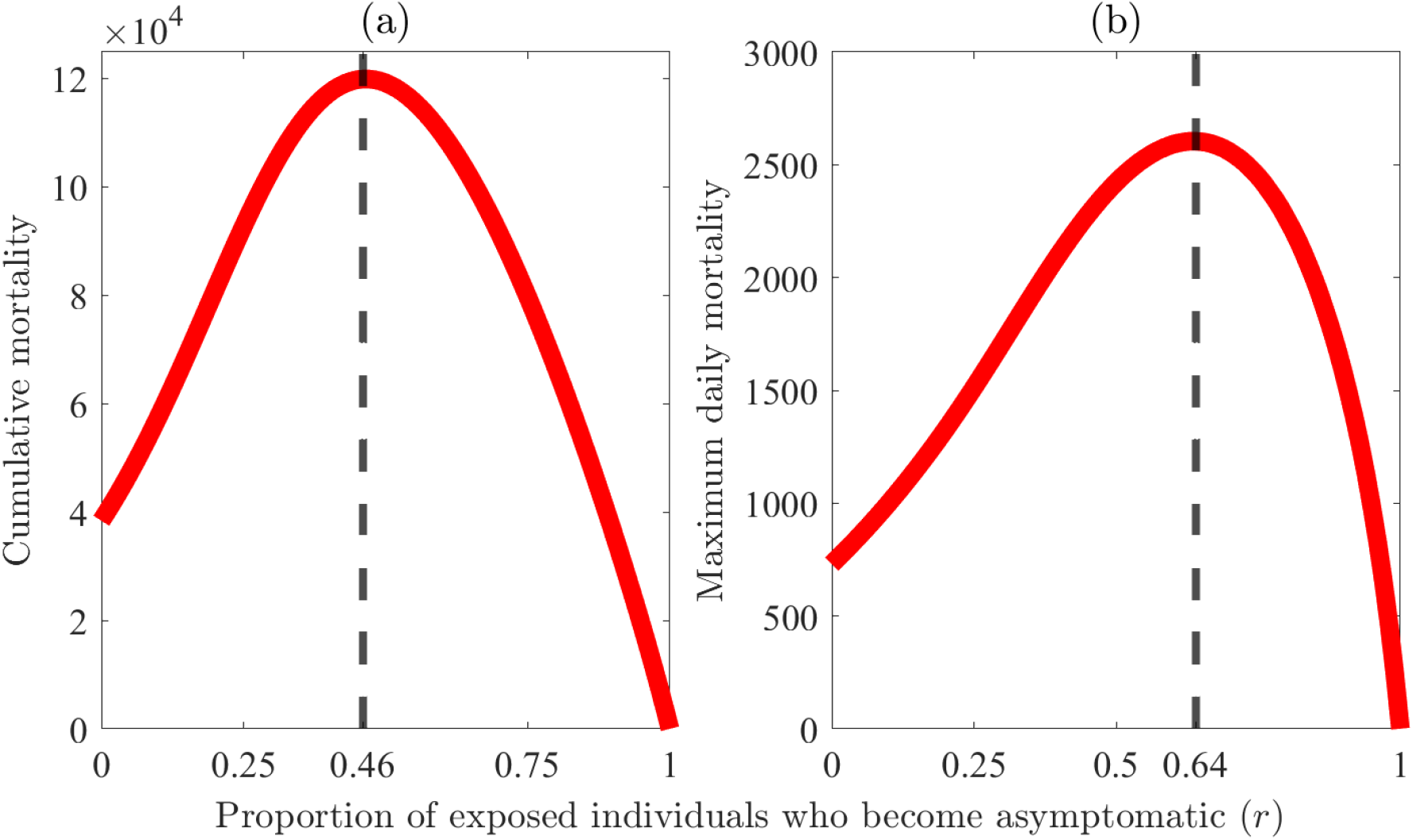
Simulations of the behavior-epidemiology model (2.4) assessing the impact of change in the proportion of exposed individuals who become asymptomatic at the end of the exposed period (*r*) on mortality. The change in (a) cumulative mortality and (b) maximum daily death is displayed for varying values of *r*. The vertical dashed line in (a) depicts the value of *r* that maximizes cumulative mortality while in (b) it depicts a value of *r* that maximizes daily mortality. The figures are generated by varying the value of *r* between 0 and 1 in the behavior-epidemiology model (2.4) while taking the remaining parameter values from Tables 3 and 4.

#### 3.3.3 Impact of the proportion of exposed individuals who become asymptomatically-infectious at the end of exposed period (*r*) on human behavior

Finally, the behavior-epidemiology model (2.4) is simulated to assess the impact of the change in the level of the proportion of exposed individuals who become asymptomatic at the end of the exposed period (*r*) on inducing human behavior changes during the first wave of the pandemic in the United States. Specifically, this is assessed by generating the profiles of the number of susceptible individuals in the adherent group (i.e., *S*_2_(*t*)) for various values of *r* between 0 and 1. Furthermore, these simulations are carried out for various values of the modification parameter for the infectiousness of asymptomatically-infectious individuals, in relation to the infectiousness of symptomatic individuals (*η*_*a*_) to assess whether or not the infectiousness of asymptomatic individuals also affects the impact of *r* on inducing the changes in behavior during the first wave. The results obtained, for the dynamics of the number of susceptible adherent individuals (*S*_2_), as a function of *r* and for various values of *η*_*a*_, are depicted in Figure 8. This figure shows, first of all, that the profile of *S*_2_ decreases with increasing values of *r* (i.e., as the proportion of exposed individuals who become asymptomatic at the end of the exposed period increases, the impact of behavior change to induce transition from the non-adherent susceptible group, *S*_1_, to the adherent susceptible group, *S*_2_, decreases; hence, the number of individuals in the *S*_2_ class decreases). Furthermore, for decreasing values of *r* ∈ [0, 0.8], the population of adherent susceptible individuals (*S*_2_) increases from near zero, rises to a peak and essentially reaches a stable positive equilibrium value for all values of *η*_*a*_ used in the simulations. Similarly, while the peak and positive equilibrium values of *S*_2_ corresponding to the case where *r* was chosen to be below its baseline value of *r* = 0.4 (i.e., *r* = 0 or *r* = 0.2) are always above those corresponding to the baseline scenario (compare the green and gold curves in Figure 8 with the blue curves for the baseline scenario with *r* = 0.4). Conversely, for cases where *r* exceeds the baseline value of *r* = 0.4 (i.e., *r* = 0.6 and *r* = 0.8), the peak and positive equilibrium values of *S*_2_ are consistently lower than the baseline profile compare the magenta and black curves in Figure 8 with the blue curve). The profile of *S*_2_ is the highest when *r* = 0, regardless of the value of *η*_*a*_. In other words, behavior change that induces the transition from the non-adherent susceptible group (*S*_1_) to the corresponding adherent susceptible group (*S*_2_) is the highest when all exposed individuals become symptomatic at the end of the exposed period (this is intuitive since the presence of a large number of symptomatic individuals in the community is highly likely to induce behavior change from the non-adherent susceptible class, *S*_1_, to the adherent susceptible class, *S*_2_). It should also be mentioned that the increase or decrease in the values of *S*_2_ at the peak and at the end of the first wave, in relation to the corresponding values for the baseline scenario (value of *r* = 0.4), become more pronounced as the value of *η*_*a*_ increases from values below one to values through and above one. For instance, for the case with *r* = *η*_*a*_ = 0.8, the values of *S*_2_ at the peak and at the end of the first wave were 2.56*×* 10^8^ and 2.44 *×* 10^8^, respectively. These values correspond to a 12.62% and 12.86% decrease from the corresponding values obtained for the baseline value of *r* = 0.4, respectively (compare the black and the blue curves in Figure 8(a)). On the other hand, the values of *S*_2_ at the peak and at the end of the first wave corresponding to the case with *r* = 0.8 and *η*_*a*_ = 2.275 were 1.91*×* 10^8^ and 5.98 *×* 10^7^, which correspond to a reduction of 30.04% and 64.40% from the respective baseline values (compare the black and blue curves in Figure 8(d)). It is also worth noting from Figure 8 that the profile of *S*_2_ becomes negligible whenever *r* = 1 (see red curves in Figure 8), regardless of the value of *η*_*a*_. For instance, it can be seen from the red curve in Figure 8(d), for the case where *η*_*a*_ = 2.275 and *r* = 1 (i.e., all exposed individuals become asymptomatic at the end of the exposed period), that the equilibrium value of *S*_2_ is about 2,550. In other words, behavior change that induces transition from the non-adherent susceptible group (*S*_1_) to the adherent susceptible group (*S*_2_) is minimized when all exposed individuals become asymptomatic at the end of the exposed period (this is also intuitive considering the fact that when all exposed individuals remain asymptomatic at the end of the exposed period, non-adherent susceptible individuals are highly unlikely to change their behavior and become adherent because they do not see people with symptoms of the SARS-CoV-2 pandemic in the community).

**Figure 8:**
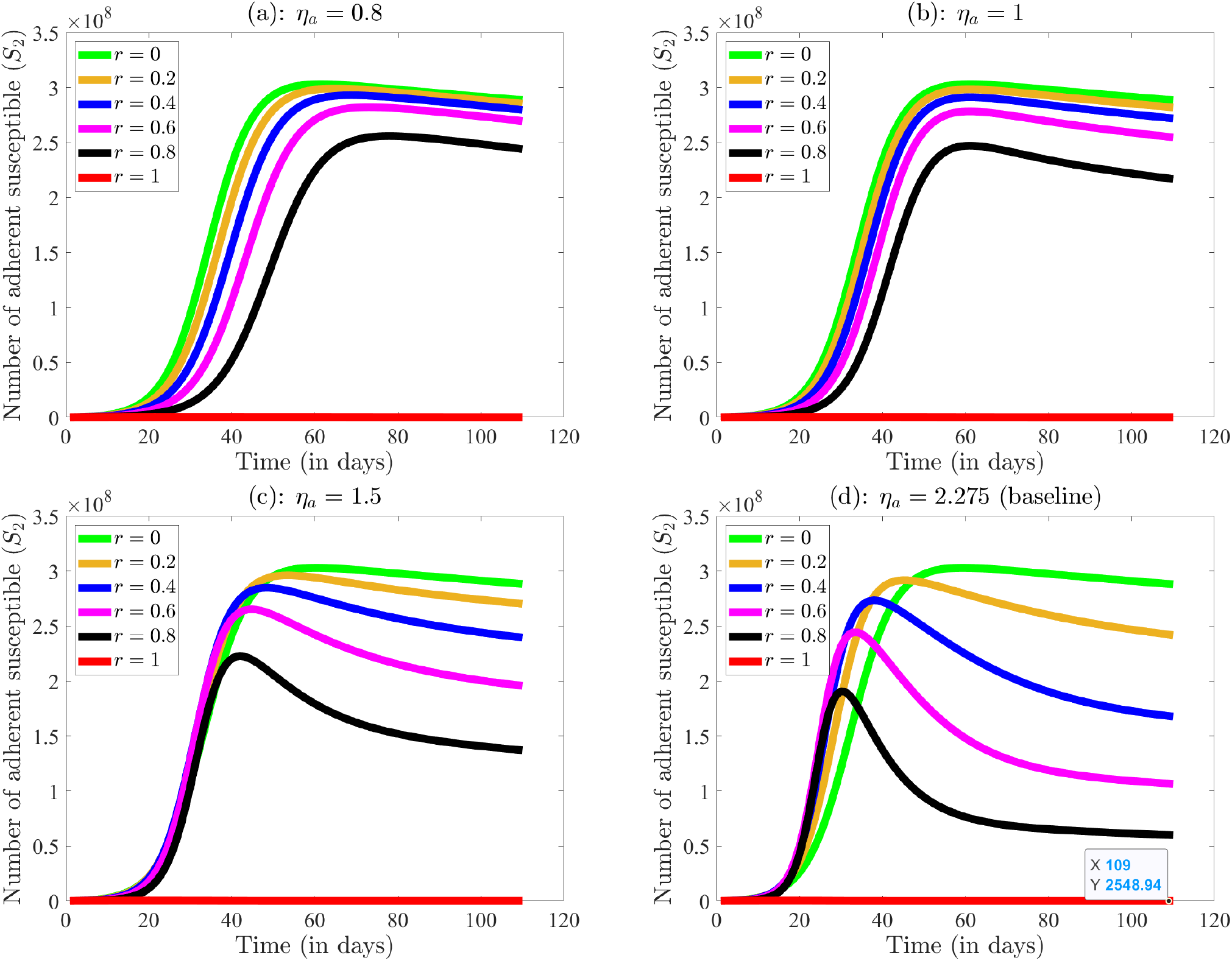
Simulations of the behavior-epidemiology model (2.4) assessing the impact of change in the proportion of exposed individuals who become asymptomatic at the end of the exposed period (*r*) on human behavior. The impact of *r* on human behavior is assessed by plotting the number of susceptible individuals in group 2 (*S*_2_) for several values of *r* (specifically, *r* = 0, *r* = 0.2, *r* = 0.4, *r* = 0.6, *r* = 0.8 and *r* = 1) for (a) *η*_*a*_ = 0.8, (b) *η*_*a*_ = 1, (c) *η*_*a*_ = 1.5 and *η*_*a*_ = 2.275. Figure (d) (corresponding to *η*_*a*_ = 2.275) shows that the equilibrium of *S*_2_ curve when *r* = 1 is minute (approximately 2, 549 people) but non-zero. The figures are generated using the behavior-epidemiology model (2.4) with aforementioned values of *r* and *η*_*a*_ while keeping the remaining parameter values as in Tables 3 and 4.

It should be noted from Figure 8 that for the case where all exposed individuals become asymptomatically-infectious at the end of the exposed period (i.e., *r* = 0), the profile of *S*_2_ is independent of the value of the parameter *η*_*a*_ (for the relative infectiousness of asymptomatically-infectious, in comparison to symptomatic infectious individuals; that is, the profile of *S*_2_, shown by the green curves in Figure 8, are all the same). This result is intuitive considering the fact that, for the case *r* = 0, all exposed individuals become symptomatic (hence, there is no asymptomatic transmission in the community, thereby negating the impact of *η*_*a*_, a parameter associated with the transmissibility of asymptomatic infectious individuals). It can also be seen from Figure 8 that, for values of *r* chosen in the range *r* ∈ [0.2, 0.8], an increase in the value of the parameter *η*_*a*_ causes the corresponding *S*_2_ profile to peak sooner (by shifting the curves to the left), in addition to decreasing the size of the peak attained and decreasing the equilibrium value of *S*_2_ (for example, compare the black curves in all four subplots of Figure 8 with increasing *η*_*a*_ value). This phenomenon is observed because an increase in the *η*_*a*_ corresponds to an increase in the relative contribution of asymptomatic infectious individuals in spreading the disease and, thus, an increase in the control reproduction number (ℛ_*C*_) for *r* ∈ (0, 1]. For example, for the baseline value of *r* = 0.4 (depicted by the blue curves in all four subplots of Figure 8) and *η*_*a*_ set at 0.8, 1, 1.5 and 2.275 (while the remaining parameters of the behavior-epidemiology are kept at their baseline values in Tables 4 and 3), the control reproduction number (ℛ_*C*_) of the model is computed to be 1.45, 1.60, 1.90 and 2.42, respectively.

Similar dynamics were observed for the profiles of *S*_1_ for various values of *r* and *η*_*a*_, as depicted in Figure 9. Here, the profiles of *S*_1_ plotted for the case of all infected individuals being symptomatic (i.e., *r* = 0) are identical. When asymptomatically-infectious individuals transmit at a rate lower than symptomatically-infectious individuals (i.e., when *η*_*a*_ *<* 1, such as the case with *η*_*a*_ = 0.8 *<* 1, as depicted in Figure 9(a)), an increase in the proportion of exposed individuals who become asymptomatically-infectious at the end of the exposed period (*r*) decreases the control reproduction number (hence, decreases the disease incidence). In other words, an increase in *r* for *η*_*a*_ *<* 1 leads to a slower decrease in *S*_1_. On the other hand, if *η*_*a*_ *>* 1 (as observed in Figures 9(c) and (d)), the control reproduction number increases with increasing values of *r* (and this causes a sharper decrease in the profile of *S*_1_ as the value of *r* increases). For instance, for the baseline value of *η*_*a*_ = 2.275 and *r* chosen to be 0, 0.2, 0.4, 0.6, 0.8 and 1 (as depicted in Figure 9(d)), the control reproduction number of the behavior-epidemiology model takes the value 1.58, 2.00, 2.42, 2.84, 3.26 and 3.68, respectively. A contour plot of the control reproduction number (ℛ_*C*_) of the behavior-epidemiology model, as a function of *r* and *η*_*a*_, is depicted in Figure C.2 of Appendix C. This figure shows that the control reproduction number increases (decreases) with increasing value of *r* if *η*_*a*_ *>* 1 (*η*_*a*_ *<* 1). For the case of *r* = 0, this figure shows a control reproduction of 1.58 for any value of *η*_*a*_.

**Figure 9:**
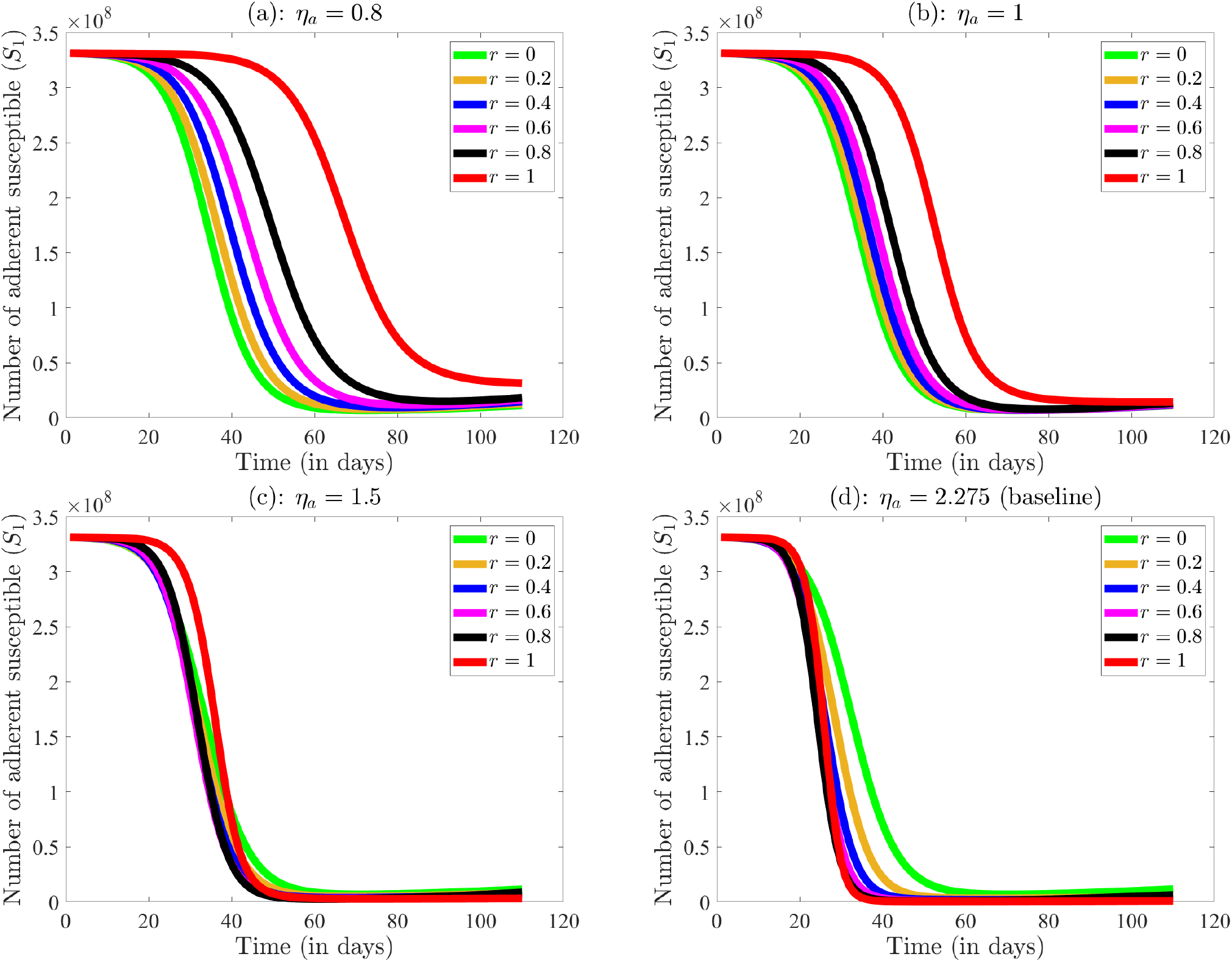
Simulations of the behavior-epidemiology model (2.4) assessing the impact of change in the proportion of exposed individuals who become asymptomatic at the end of the exposed period (*r*) on human behavior. The impact of *r* on human behavior is assessed by plotting the number of susceptible individuals in group 1 (*S*_1_) for several values of *r* (specifically, *r* = 0, *r* = 0.2, *r* = 0.4, *r* = 0.6, *r* = 0.8 and *r* = 1) for (a) *η*_*a*_ = 0.8, (b) *η*_*a*_ = 1, (c) *η*_*a*_ = 1.5 and *η*_*a*_ = 2.275. The figures are generated using the behavior-epidemiology model (2.4) with aforementioned values of *r* and *η*_*a*_ while keeping the remaining parameter values as in Tables 3 and 4.

## 4 Discussion and Conclusion

This study presents a novel mathematical model for the transmission dynamics and control of the SARS-CoV-2 pandemic in the United States that explicitly incorporates the impacts of changing human behavior. Specifically, the model was formulated by stratifying the total population into two groups based on their adherence or lack thereof to public health interventions, and considered five metrics for human behavior, namely behavior change due to: (a) information received by members of one group from members of the other group (b) the level of symptomatic transmission in the community (c) the size of the proportion of non-symptomatic individuals in the community, (d) the level of disease-induced mortality in the community and (e) the degree of fatigue to adherence to intervention and mitigation measures (such as wearing a face mask in public, observing social distancing and community lockdowns etc.).

The resulting two-group behavior-epidemiology model, which takes the form of a relatively large deterministic system of nonlinear differential equations, was parameterized using observed cumulative SARS-CoV-2 mortality data, at the national level, for the United States during the first wave of the pandemic (March to June, 2020). The model was then rigorously analysed to gain qualitative insights into its dynamical features. Such analyses revealed that the disease-free equilibrium of the model is locally-asymptotically stable whenever a certain epidemiological quantity (known as the *control reproduction number*, denoted by ℛ_*C*_) is less than one, and unstable when the quantity exceeds one. The epidemiological consequence of this result is that the disease can be effectively controlled in the community (when ℛ_*C*_ *<* 1) if the initial number of infected individuals introduced into the community is small enough. In other words, significant outbreaks will not occur if the intervention and mitigation measures implemented can bring (and maintain) this threshold quantity to a value less than one. The model was then simulated to quantify the impact of behavior changes on the trajectory and burden of the disease, in addition to assessing the population-level impact of public health intervention and mitigation measures (specifically, basic nonpharmaceutical interventions, such as the use of face mask by individuals in the adherent group).

As stated above, the two-group behavior-epidemiology model was fitted and cross-validated using the observed cumulative mortality data for the United States during the first wave of the SARS-CoV-2 pandemic (which corresponds to the period from March 1, 2020, to June 18, 2020; we used the segment of the data from March 1, 2020 to May 29, 2020 for fitting the model, and the data for the remaining segment, from May 30, 2020 to June 20, 2020, for cross-validation). We showed very good fit for the cumulative mortality data (and we used the estimated parameters, together with the fixed parameters of the behavior-epidemiology model, to predict the observed daily mortality for the SARS-CoV-2 pandemic during the first wave in the United States, showing a very good prediction). We showed that, unlike for the case of the behavior-epidemiology, the behavior-free analog of the behavior-epidemiology model (i.e., the two-group model without human behavior changes explicitly incorporated into the model) did not do as well in correctly capturing the observed trajectory and burden of the pandemic during the first wave. Specifically, while the behavior-epidemiology model projected the daily mortality trend better during the fitting and cross-validation periods, the behavior-free model did not capture the correct trajectory (particularly during the early stages) of the pandemic and underestimated the observed additional mortality that occurred during the cross-validation period by about 35.49% (the behavior-epidemiology model underestimated this metric by only 12.59%). We compared the goodness of fit of the two models using various error metrics, such as sum of squared error (SSE), and showed, in all of these metrics, that the behavior-epidemiology model did far better in fitting the observed data (i.e., capturing the correct trajectory of the pandemic during the first wave) and in predicting the observed daily mortality (for the first wave), in comparison to the behavior-free model. In other words, this study clearly shows that explicitly incorporating human behavior into epidemiological models enhances their ability to correctly capture observed trends/trajectory of the pandemic, as well as enhancing their capability to make accurate prediction of the future course and burden of the pandemic. This is, perhaps, one of the earlier studies to clearly show (and quantify) the importance of explicitly including elements of human behavior into epidemiological models for the transmission dynamics and control of infectious diseases of major public health significance (such as the SARS-CoV-2 pandemic).

A major feature of the SARS-CoV-2 pandemic is the role asymptomatic infectious individuals play in generating new infections. Numerous modeling studies showed that this cohort of infectious individuals account for a sizable proportion of new cases of SARS-CoV-2 in the community, estimated to be in the range between 50% to 70% [56–58, 67–69]. Our simulations of the behavior-epidemiology and behavior-free model showed that, while the former estimated the proportion of new cases generated by asymptomatic infectious individuals to be about 70%, the latter model estimated this proportion to be about 84% (suggesting that the behavior-free model may have over-estimated the role of asymptomatic infectious individuals to generate new cases). In other words, the behavior-epidemiology model appears to do better than the behavior-free model even under this metric (of correctly quantifying the role of asymptomatic infectious individuals in generating new infections in the community).

We carried out extensive numerical simulations of the behavior-epidemiology models to determine which of the five behavior metrics considered in this study were more influential in inducing positive behavior changes (to minimize risk of acquisition and/or transmission of infection) during the first wave of the SARS-CoV-2 pandemic in the United States. The results obtained showed that the behavior change metric that induces the greatest positive behavior change (as measured in terms of the transition from the non-adherent group to the adherent group) is the behavior change due to the level of disease-induced mortality in the community, followed by behavior change due to the level of symptomatic individuals in the community (in other words, our study shows that, during the first wave, people are more likely to change behavior, and begin to strictly adhere to public health intervention and mitigation measures implemented, if they see a significant increase in mortality due to the disease in addition to a significant increase in the number of people with symptoms of the disease in the community). Our simulations also showed that behavior change induced by fatigue to interventions (i.e., wearing a mask, in this case) had only a very marginal (or essentially no) effect in inducing a negative behavior change (from adherent to non-adherent group) during the first wave of the pandemic in the United States. In other words, our study showed that the overwhelming proportion of individuals in the adherent group did not experience significant level of fatigue to intervention during the first wave of COVID-19 pandemic in the United States.

Extensive numerical simulations were also carried out to assess the impact of the the proportion of exposed individuals who become asymptomatically-infectious at the end of the exposed period (denoted by the parameter *r*) on the burden of the SARS-CoV-2 pandemic, in addition to inducing behavior change to interventions, during the first wave in the United States. We showed that, using the baseline values of the parameters of the behavior-epidemiology model in the simulations (where asymptomatic infectious individuals were at least twice more infectious than symptomatic infectious individuals), the control reproduction number (ℛ_*C*_) of the behavior-epidemiology model increases with increasing values of *r*. Thus, in this case, an increase in the proportion of exposed individuals who eventually become asymptomatically-infectious resulted in an increase in the burden of the pandemic (since an increase in ℛ_*C*_ implies increase in average number of new cases and, potentially, increase in number of hospitalizations and deaths). On the other hand, if asymptomatic infectious were not as infectious as symptomatic infectious individuals (i.e., if the parameter *η*_*a*_ for the relative infectiousness of asymptomatic individuals is less than one), while all other parameters of the model are kept at their baseline values, the control reproduction number decreases with increasing values of *r*. In other words, our study showed that the ability of asymptomatic infectious individuals to significantly increase the burden of the pandemic depends on the relative infectiousness of asymptomatic infectious individuals, in comparison to the infectiousness of symptomatic individuals. Nonetheless, this result highlights the importance of reducing the level of asymptomatic transmission in the community (through strategies such as widespread testing and rapid isolation and treatment of asymptomatic individuals).

Simulations of the behavior-epidemiology model further showed the existence of two critical threshold values of the proportion *r*, of exposed individuals who become asymptomatic infectious at the end of the exposed period, namely one that maximizes cumulative SARS-CoV-2 mortality and another that maximizes the daily SARS-CoV-2 mortality during the first wave of the pandemic in the United States. Specifically, we showed, based on the parameter values used in the simulations, the cumulative mortality is maximized if the proportion of exposed individuals who become asymptomatic at the end of the exposed period is 46%. Similarly, daily SARS-CoV-2 mortality (i.e., the peak of daily mortality during the first wave) is maximized if the proportion *r* is 64%. In summary, these results show that there is a threshold value of *r* (estimated to be 46% in our study) that maximizes the overall burden of the disease on the healthcare system during the entire duration of the first wave, and another (estimated to be 64%) that maximizes a single-day mortality burden on the healthcare system.

Finally, simulations were carried out to assess the impact of the parameter *r* (for the proportion of exposed individuals who become asymptomatic) on inducing behavior changes during the first wave of the pandemic. Specifically, simulations are carried out for various values of the modification parameter for the infectiousness of asymptomatically-infectious individuals, in relation to the infectiousness of symptomatic individuals (*η*_*a*_) to assess whether or not the infectiousness of asymptomatic individuals also affects the impact of *r* on inducing the changes in behavior during the first wave. It was shown that, regardless of the value of the parameter *η*_*a*_ used in the simulations (we considered *η*_*a*_ *<* 1, *η*_*a*_ = 1 and *η*_*a*_ *>* 1), the profile of susceptible individuals in the community who strictly adhere to public health interventions (*S*_2_) is maximized (measured in terms of the peak and equilibrium values of *S*_2_ curve) when all infectious individuals are symptomatic (i.e., *r* = 0). As *r* increases (and exposed individuals begin to become asymptomatically-infectious at the end of the exposed period), the rate at which susceptible non-adherent individuals (i.e., individuals in the *S*_1_ class) change their behavior to strictly adhere to interventions (i.e., move to the *S*_2_ class) decreases (i.e., the peak and equilibrium values of the *S*_2_ curves decrease under the scenario with increasing values of *r* from 0). When all infectious individuals are asymptomatic (i.e., *r* = 1), the profile of the adherent susceptible population (*S*_2_) is minimized (with a relatively small number of individuals in the *S*_2_ class at equilibrium). Thus, this study showed that, as the number of exposed individuals who become asymptomatically-infectious at the end of the exposed period (*r*) increases, the level or likelihood of positive behavior change by non-adherent susceptible individuals (to become adherent susceptible individuals) decreases. Thus, it can be concluded from this result that the likelihood or level of positive behavior change to public health interventions reduces for a disease where a large proportion of exposed individuals become asymptomatically-infectious at the end of the exposed period (this could be due to high level of community-wide immunity against the disease or the emergence of new variants where most infected individuals are asymptomatic, etc). Furthermore, although high values of *r* (i.e., high proportion of exposed individuals who become asymptomatically-infectious at the end of the exposed period) can increase ℛ_*C*_ (hence, increase disease incidence, particularly if the effective contact rate of asymptomatically-infectious individuals exceed that of symptomatically-infectious individuals), it can also decrease the level of positive behavior change (from non-adherent to adherent) as well as reduce disease severity, hospitalization and disease-induced mortality in the community. Thus, as more new infected individuals become asymptomatically-infectious, the level of positive behavior change, as well as disease severity, hospitalization and disease-induced mortality in the community can be expected to significantly decrease (while new cases may rise, particularly if asymptomatic-infectious individuals have higher contact rate, in comparison to symptomatic infectious individuals).

Some of the limitations of this modeling study include: the behavior-epidemiology model assumes that behavior change from one group to the other occurs only due to the five metrics we enumerated (i.e., contact with individuals from a different group, the level of symptomatic transmission in the community, the proportion of non-symptomatic individuals in the community, the level of reported disease-induced mortality in the community, and the level of intervention fatigue in the community). However, there may be various other metrics that could induce behavior change during a pandemic like COVID-19 (for example, adherence could be forced upon a community by the government). Moreover, the model does not explicitly account for individuals who will never adhere to intervention due to socio-economic reasons (e.g., the case of some meat factory workers during the SARS-CoV-2 pandemic [70, 71]) or political polarization of the community (where some individuals within segments of the community will simply deny the existence of the disease, let alone adhere to intervention measures against it, due to their political or other leanings). The latter limitation could be addressed by extending the two-group model to a three-group model, where the additional group is for individuals in the community who never change their non-adherent status.

Future work should explore fitting the proposed model to other time periods and finer spatial resolutions, such as state and city levels. In addition, we envision that data from surveys, where participants are queried in different time periods of the pandemic about their daily habits regarding contacts with other individuals outside their household (i.e., people within their social network), going to the office or places of worship, using face masks, or their commuting choices (e.g., taking public transport), could help generate data that can be used to improve the estimate of some of the parameters in the behavior metrics introduced in this paper [72].

## Data Availability

Publicly-available data was used in the study, and all relevant references/sources are provided.

https://coronavirus.jhu.edu/map.html

## Acknowledgments

ABG acknowledges the support, in part, of the National Science Foundation (Grant Number: DMS-2052363; transferred to DMS-2330801). SS acknowledges the support of the Fulbright Foreign Student Program.

**Appendix A Equations for the Behavior-free Model**

The behavior-free model, discussed in Section 2.3 and obtained by setting behavior-related parameters of the model (2.4) to zero (i.e., consider the model (2.4) with 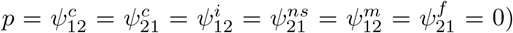, is given by the following system of nonlinear differential equations:

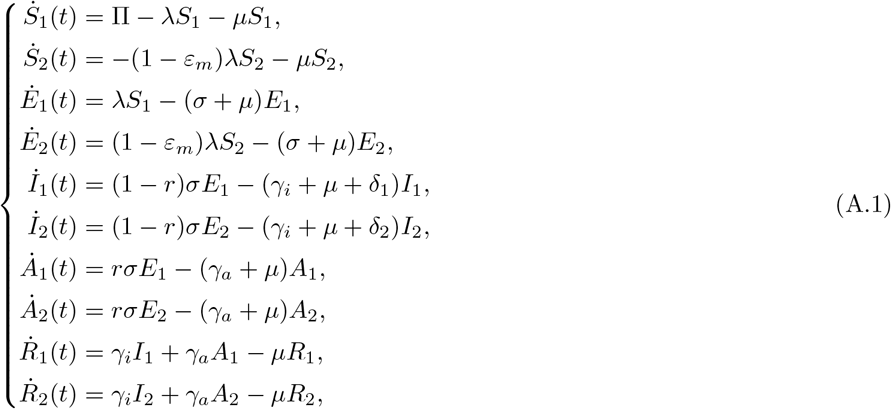

where *λ* (i.e., the *force of infection*) for the behavior-free model (A.1) is as given in (2.3)). Furthermore, like for the case of the behavior-epidemiology model, the state variables and parameters of the behavior-free model are described in Tables 1 and 2.

**Appendix B Statistical Metrics for Goodness of Fit for the Models: SSE, RMSE, RMSLE and Correlation Coefficients**

The comparison of goodness of fit of the behavior-epidemiology (2.4) and behavior-free (A.1) models during the fitting period (i.e., March 1, 2020, to May 29, 2020) and during the cross-validation period (i.e., May 30, 2020, to June 18, 2020) was made in Section 2.3.1 using SSE. It was observed that the behavior-epidemiology (2.4) has an overall lower SSE. In this Appendix, we provide some additional metrics to compare the goodness of fit of the two models.

Root mean squared error (RMSE) provides an average difference between observed and model-predicted values. The RMSE can be expressed as a square root of the mean of SSE (known as MSE) [73, 74]:

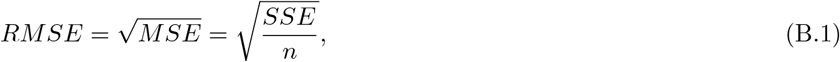

where SSE is as defined in (2.15) and *n* is the total number of observations. Since the observation (especially cumulative mortality) is large, it is also reasonable to consider root mean squared logarithmic error (RMSLE), which involves taking the logarithm of predicted and observed values. Specifically, the RMSLE is expressed as [75]

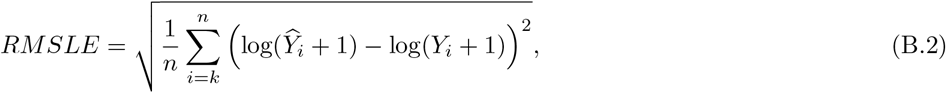

where *Y*_*i*_ be the observed data on day *i*, 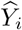 is the model’s output data on day *i, k* is the starting point of the sum and *n* is the total number of the given or available (observed) data points. We note that, similar to SSE, models with smaller values of RMSE and RMSLE may be preferable. In particular, when these metrics yield a value of zero, it indicates no discrepancy between the observed and predicted data —signifying a perfect prediction by the model. The SSE, RMSE, and RMSLE values for both the behavior-epidemiology and behavior-free models during the data fitting p eriod a re p resented i n Table B.1, while those during the cross-validation period are given in Table B.2.

**Table B.1:** The sum squared error (SSE), root mean squared error (RMSE) and root mean squared logarithmic error (RMSLE) on observed cumulative and daily mortality data with respective projection from behavior-epidemiology model (2.4) and behavior-free model (A.1) during data fitting period (March 1, 2020 to May 29, 2020).

|  | Behavior-epidemiology model | Behavior-free model |
| --- | --- | --- |
| SSE on cumulative mortality | $4.99 \times 10^7$ | $7.81 \times 10^8$ |
| SSE on daily mortality | $4.87 \times 10^6$ | $1.31 \times 10^7$ |
| RMSE on cumulative mortality | $7.45 \times 10^2$ | $2.95 \times 10^3$ |
| RMSE on daily mortality | $2.33 \times 10^2$ | $3.81 \times 10^2$ |
| RMSLE on cumulative mortality | 0.28 | 1.53 |
| RMSLE on daily mortality | 0.34 | 1.42 |

**Table B.2:** The sum squared error (SSE), root mean squared error (RMSE) and root mean squared logarithmic error (RMSLE) on observed cumulative and daily mortality data with respective projection from behavior-epidemiology model (2.4) and behavior-free model (A.1) during data cross-validation period (May 30, 2020 to June 18, 2020).

|  | Behavior-epidemiology model | Behavior-free model |
| --- | --- | --- |
| SSE on cumulative mortality | $1.49 \times 10^7$ | $1.36 \times 10^8$ |
| SSE on daily mortality | $9.02 \times 10^5$ | $3.12 \times 10^6$ |
| RMSE on cumulative mortality | $8.64 \times 10^2$ | $2.60 \times 10^3$ |
| RMSE on daily mortality | $2.12 \times 10^2$ | $3.95 \times 10^2$ |
| RMSLE on cumulative mortality | 0.007 | 0.022 |
| RMSLE on daily mortality | 0.33 | 0.80 |

**Figure B.1:**
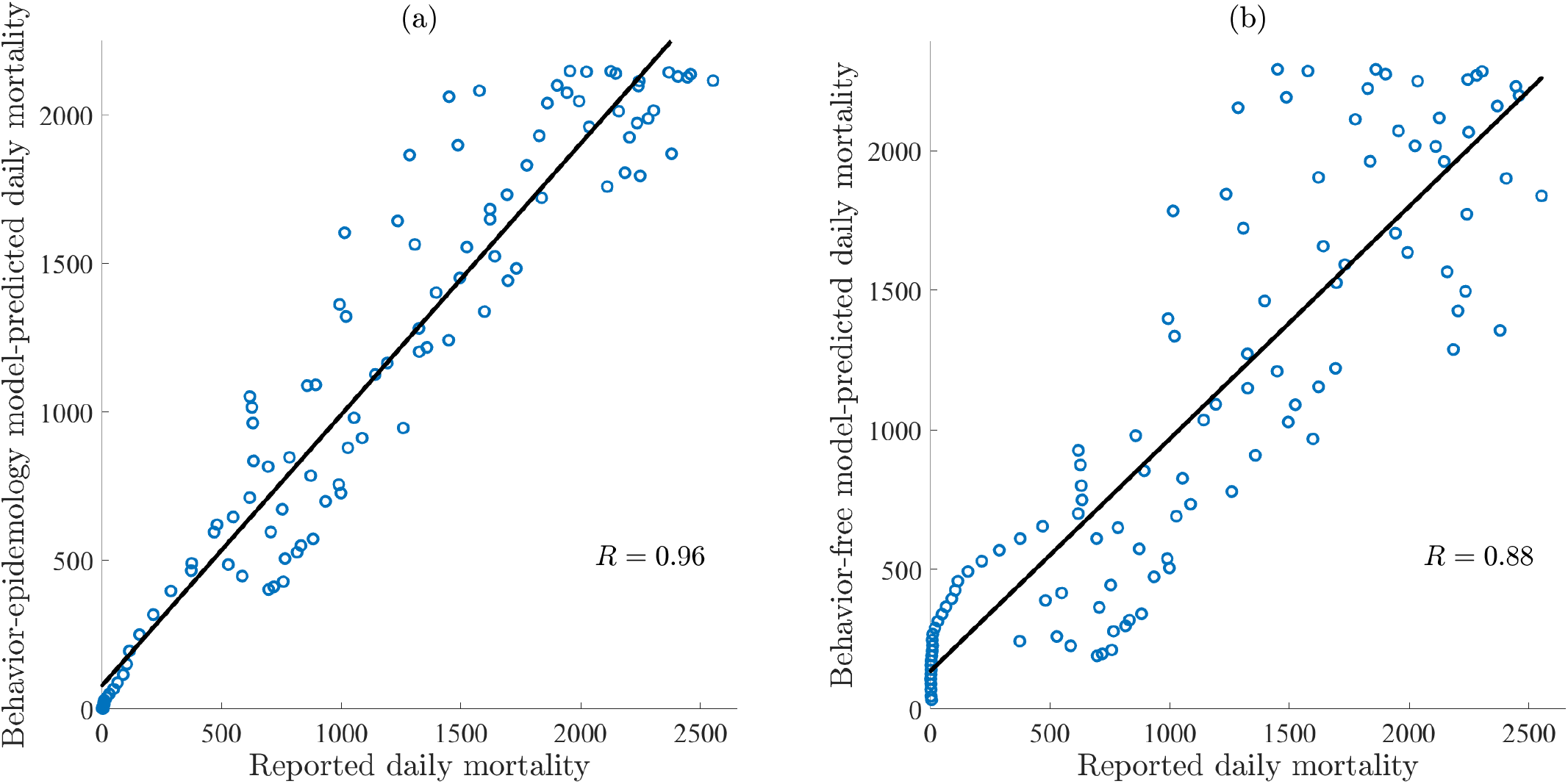
Correlation between reported daily mortality and predicted daily mortality with (a) behavior-epidemiology model (2.4) and (b) behavior-free model (2.12). The data points are given by blue circles while the black line represents the best linear fit to the data. The correlation coefficient (*R*) for behavior-epidemiology and behavior-free models are 0.96 and 0.88, respectively.

In Figure B.1, the reported daily mortality is plotted with predicted daily mortality obtained from the behavior-epidemiology model (2.4) (Figure B.1(a)) and behavior-free model (2.12) (Figure B.1(b)) for the entire duration of the simulation. It is observed that the correlation coefficients (*R*) and r-squared (*R*^2^) [60] associated with the behavior-epidemiology model (*R* = 0.96 and *R*^2^ = 0.92) are closer to one than that from the behavior-free model (*R* = 0.88 and *R*^2^ = 0.77), thus signifying that the behavior-epidemiology model more accurately captures the daily mortality trend.

**Appendix C Miscellaneous Figures**

**Figure C.1:**
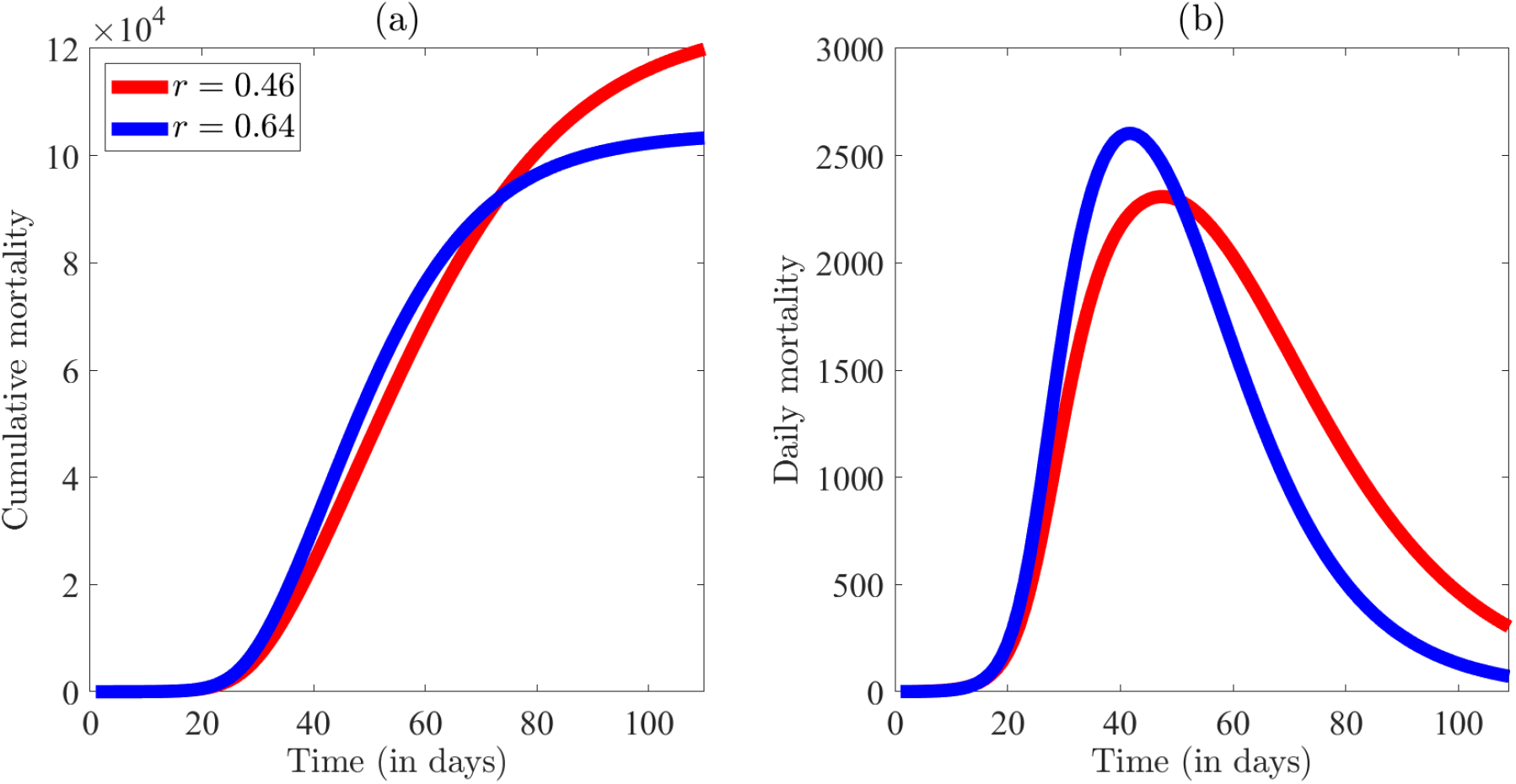
Simulation of the behavior-epidemiology model (2.4) depicting the impact of two specific values of the proportion of newly infected individuals who become asymptomatically-infectious at the end of the exposed period (*r*) on mortality. The (a) cumulative mortality and (b) daily mortality figures are generated for values of *r* = 0.46 and *r* = 0.64 with remaining parameters as in Tables 3 and 4.

**Figure C.2:**
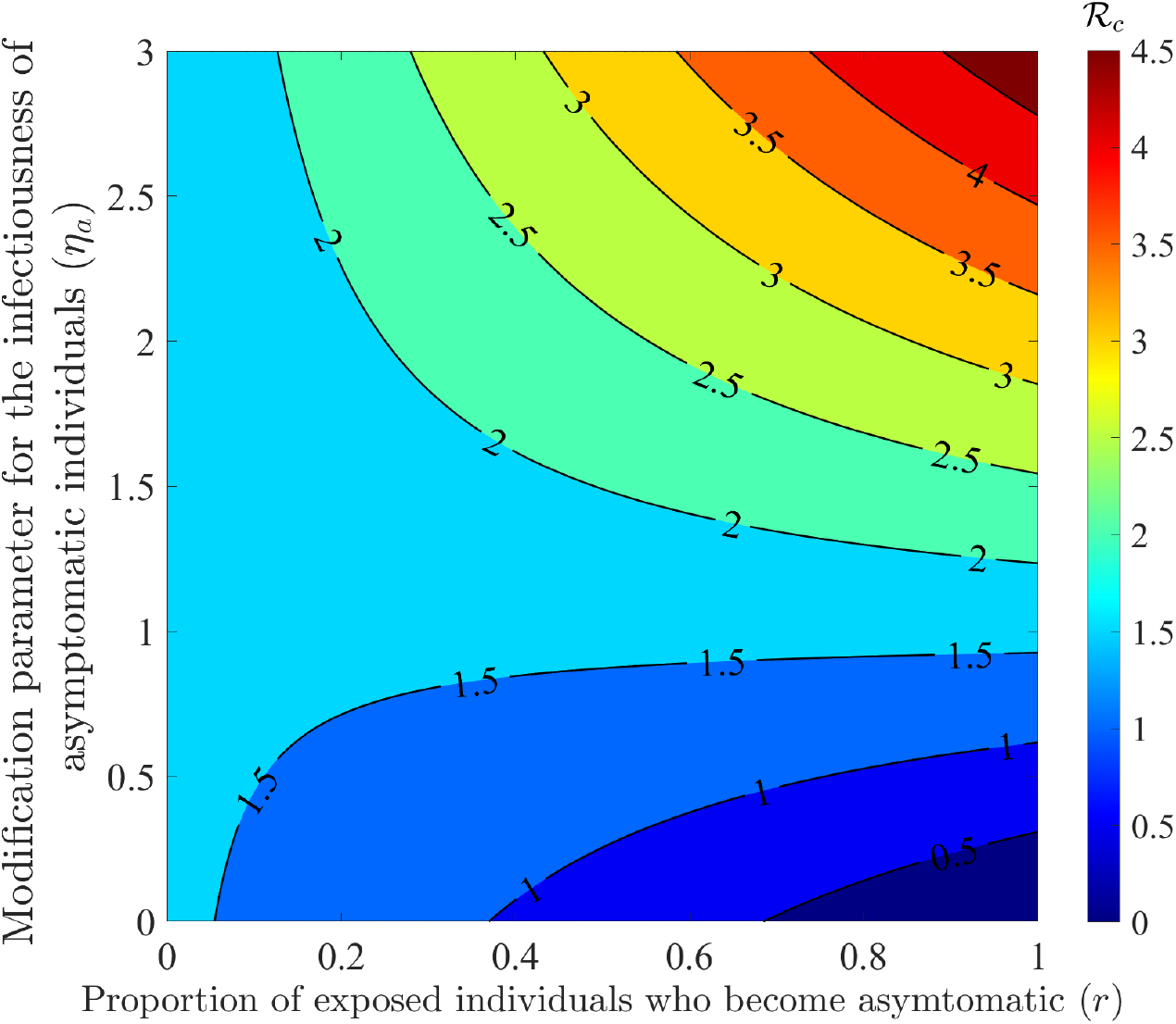
Contour plot generated using the behavior-epidemiology model (2.4) depicting the impact of of the proportion of exposed individuals who become asymptomatic at the end of the exposed period (*r*) and the modification parameter for the infectiousness of asymptomatic individuals (*η*_*a*_) on the control reproduction number (*ℛ*_*C*_).

## References

[1] N. LePan, N. Routley, and H. Schell, “Visualizing the history of pandemics.” https://www.visualcapitalist.com/history-of-pandemics-deadliest/, 2020. Accessed September 18, 2023.

[2] D. Huremović, “Brief history of pandemics (pandemics throughout history),” Psychiatry of Pandemics: a Mental Health Response to Infection Outbreak, pp. 7–35, 2019.

[3] J. Piret and G. Boivin, “Pandemics throughout history,” Frontiers in Microbiology, vol. 11, p. 631736, 2021.

[4] World Health Organization, “WHO coronavirus disease (COVID-19) dashboard.” https://covid19.who.int. Accessed September 17, 2023.

[5] A. Shalal, “IMF sees cost of covid pandemic rising beyond $12.5 trillion estimate.” https://www.reuters.com/business/imf-sees-cost-covid-pandemic-rising-beyond-125-trillion-estimate-2022-01-20/, 2022.

[6] R. M. Barber, R. J. Sorensen, D. M. Pigott, C. Bisignano, A. Carter, J. O. Amlag, J. K. Collins, C. Abbafati, C. Adolph, A. Allorant, et al., “Estimating global, regional, and national daily and cumulative infections with sars-cov-2 through nov 14, 2021: a statistical analysis,” The Lancet, vol. 399, no. 10344, pp. 2351–2380, 2022.

[7] C. for Disease Control, Prevention, et al., “Nearly one in five American adults who have had COVID-19 still have “long COVID”.” https://www.cdc.gov/nchs/pressroom/nchs_press_releases/2022/20220622.htm, 2022.

[8] F. Luo, R. Ghanei Gheshlagh, S. Dalvand, S. Saedmoucheshi, and Q. Li, “Systematic review and meta-analysis of fear of COVID-19,” Frontiers in Psychology, vol. 12, p. 661078, 2021.

[9] K. M. Fitzpatrick, C. Harris, and G. Drawve, “Fear of COVID-19 and the mental health consequences in America.,” Psychological Trauma: Theory, Research, Practice, and Policy, vol. 12, no. S1, p. S17, 2020.

[10] X. Liu, W.-T. Luo, Y. Li, C.-N. Li, Z.-S. Hong, H.-L. Chen, F. Xiao, and J.-Y. Xia, “Psychological status and behavior changes of the public during the COVID-19 epidemic in China,” Infectious Diseases of Poverty, vol. 9, no. 03, pp. 20–30, 2020.

[11] D. K. Ahorsu, C.-Y. Lin, V. Imani, M. Saffari, M. D. Griffiths, and A. H. Pakpour, “The fear of COVID-19 scale: development and initial validation,” International Journal of Mental Health and Addiction, pp. 1–9, 2020.

[12] J. Roozenbeek, C. R. Schneider, S. Dryhurst, J. Kerr, A. L. Freeman, G. Recchia, A. M. Van--Der--Bles, and S. Van--Der--Linden, “Susceptibility to misinformation about COVID-19 around the world,” Royal Society Open Science, vol. 7, no. 10, p. 201199, 2020.

[13] J. Y. Cuan-Baltazar, M. J. Muñoz-Perez, C. Robledo-Vega, M. F. Pérez-Zepeda, and E. Soto-Vega, “Misinformation of COVID-19 on the internet: infodemiology study,” JMIR Public Health and Surveillance, vol. 6, no. 2, p. e18444, 2020.

[14] N. Perra, D. Balcan, B. Gonçalves, and A. Vespignani, “Towards a characterization of behavior-disease models,” PloS one, vol. 6, no. 8, p. e23084, 2011.

[15] I. Z. Kiss, J. Cassell, M. Recker, and P. L. Simon, “The impact of information transmission on epidemic outbreaks,” Mathematical Biosciences, vol. 225, no. 1, pp. 1–10, 2010.

[16] M. M. Tanaka, J. Kumm, and M. W. Feldman, “Coevolution of pathogens and cultural practices: a new look at behavioral heterogeneity in epidemics,” Theoretical Population Biology, vol. 62, no. 2, pp. 111–119, 2002.

[17] S. Funk, E. Gilad, and V. A. Jansen, “Endemic disease, awareness, and local behavioural response,” Journal of Theoretical Biology, vol. 264, no. 2, pp. 501–509, 2010.

[18] K. Yan, “Modeling the effect of human behavior on disease transmission,” 2022.

[19] S. Del Valle, H. Hethcote, J. M. Hyman, and C. Castillo-Chavez, “Effects of behavioral changes in a smallpox attack model,” Mathematical Biosciences, vol. 195, no. 2, pp. 228–251, 2005.

[20] A. G. Kummer, J. Zhang, M. Litvinova, A. Vespignani, H. Yu, and M. Ajelli, “Measuring the seasonality of human contact patterns and its implications for the spread of respiratory infectious diseases,” medRxiv, pp. 2022–02, 2022.

[21] A. d’Onofrio, P. Manfredi, and E. Salinelli, “Vaccinating behaviour, information, and the dynamics of sir vaccine preventable diseases,” Theoretical Population Biology, vol. 71, no. 3, pp. 301–317, 2007.

[22] P. Manfredi and A. D’Onofrio, Modeling the interplay between human behavior and the spread of infectious diseases. Springer Science & Business Media, 2013.

[23] F. C. Coelho and C. T. Codeço, “Dynamic modeling of vaccinating behavior as a function of individual beliefs,” PLoS Computational Biology, vol. 5, no. 7, p. e1000425, 2009.

[24] J. de Mooij, P. Bhattacharya, D. Dell’Anna, M. Dastani, B. Logan, and S. Swarup, “A framework for modeling human behavior in large-scale agent-based epidemic simulations,” Simulation, p. 00375497231184898, 2023.

[25] S. Y. D. Valle, S. M. Mniszewski, and J. M. Hyman, “Modeling the impact of behavior changes on the spread of pandemic influenza,” Modeling the interplay between human behavior and the spread of infectious diseases, pp. 59–77, 2013.

[26] Z. Wang, M. A. Andrews, Z.-X. Wu, L. Wang, and C. T. Bauch, “Coupled disease–behavior dynamics on complex networks: A review,” Physics of Life Reviews, vol. 15, pp. 1–29, 2015.

[27] L. Mao and Y. Yang, “Coupling infectious diseases, human preventive behavior, and networks–a conceptual framework for epidemic modeling,” Social Science & Medicine, vol. 74, no. 2, pp. 167–175, 2012.

[28] K. Frieswijk, L. Zino, M. Ye, A. Rizzo, and M. Cao, “A mean-field analysis of a network behavioral–epidemic model,” IEEE Control Systems Letters, vol. 6, pp. 2533–2538, 2022.

[29] H. V. Huff and A. Singh, “Asymptomatic transmission during the coronavirus disease 2019 pandemic and implications for public health strategies,” Clinical Infectious Diseases, vol. 71, no. 10, pp. 2752–2756, 2020.

[30] L. A. Nikolai, C. G. Meyer, P. G. Kremsner, and T. P. Velavan, “Asymptomatic SARS Coronavirus 2 infection: Invisible yet invincible,” International Journal of Infectious Diseases, vol. 100, pp. 112–116, 2020.

[31] C. N. Ngonghala, E. A. Iboi, and A. B. Gumel, “Could masks curtail the post-lockdown resurgence of COVID-19 in the us?,” Mathematical Biosciences, vol. 329, p. 108452, 2020.

[32] B. Espinoza, M. Marathe, S. Swarup, and M. Thakur, “Asymptomatic individuals can increase the final epidemic size under adaptive human behavior,” Scientific Reports, vol. 11, no. 1, p. 19744, 2021.

[33] D. He, J. Dushoff, T. Day, J. Ma, and D. J. Earn, “Inferring the causes of the three waves of the 1918 influenza pandemic in England and Wales,” Proceedings of the Royal Society B: Biological Sciences, vol. 280, no. 1766, p. 20131345, 2013.

[34] W. C. Roda, M. B. Varughese, D. Han, and M. Y. Li, “Why is it difficult to accurately predict the COVID-19 epidemic?,” Infectious Disease Modelling, vol. 5, pp. 271–281, 2020.

[35] F. Sayers, “Why were Denmark’s Covid models better than England’s?.” https://unherd.com/thepost/why-were-denmarks-covid-models-better-than-englands/. Accessed January 19, 2024.

[36] S. J. Brozak, B. Pant, S. Safdar, and A. B. Gumel, “Dynamics of COVID-19 pandemic in India and Pakistan: A metapopulation modelling approach,” Infectious Disease Modelling, vol. 6, pp. 1173–1201, 2021.

[37] C. N. Ngonghala, H. B. Taboe, S. Safdar, and A. B. Gumel, “Unraveling the dynamics of the Omicron and Delta variants of the 2019 coronavirus in the presence of vaccination, mask usage, and antiviral treatment,” Applied Mathematical Modelling, vol. 114, pp. 447–465, 2023.

[38] H. Howard and L. Andrews, “Another doomsday SAGE prediction that was wrong: Expert admits forecasting 6,000 Omicron deaths a day when it only reached 306 were wildly wrong because they failed to predict britons would change their behaviour.” https://www.dailymail.co.uk/news/article-10571661/SAGE-expert-says-wildly-wrong-Omicron-death-predictions-failed-account-behaviour-change.html. Accessed January 19, 2024.

[39] S. E. Eikenberry, M. Mancuso, E. Iboi, T. Phan, K. Eikenberry, Y. Kuang, E. Kostelich, and A. B. Gumel, “To mask or not to mask: Modeling the potential for face mask use by the general public to curtail the COVID-19 pandemic,” Infectious Disease Modelling, vol. 5, pp. 293–308, 2020.

[40] V. S. Ivlev, “Experimental ecology of the feeding of fishes,” Yale University Press, 1961.

[41] M. E. Bouton, “Why behavior change is difficult to sustain,” Preventive Medicine, vol. 68, pp. 29–36, 2014.

[42] B. Epstein and D. Lofquist, “US Census Bureau Today delivers state population totals for congressional apportionment. US Census Bureau.” https://www.census.gov/library/stories/2021/04/2020-census-data-release.html. Accessed December 9, 2023.

[43] Arias, Elizabeth and Tejada-Vera, Betzaida and Ahmad, Farida, “Provisional life expectancy estimates for January through June, 2020.” https://www.cdc.gov/coronavirus/2019-ncov/hcp/planning-scenarios.html, Feb. 2020. Accessed December 9, 2023.

[44] R. Li, S. Pei, B. Chen, Y. Song, T. Zhang, W. Yang, and J. Shaman, “Substantial undocumented infection facilitates the rapid dissemination of novel coronavirus (SARS-CoV-2),” Science, vol. 368, no. 6490, pp. 489–493, 2020.

[45] Q. Ma, J. Liu, Q. Liu, L. Kang, R. Liu, W. Jing, Y. Wu, and M. Liu, “Global percentage of asymptomatic SARS-CoV-2 infections among the tested population and individuals with confirmed COVID-19 diagnosis: a systematic review and meta-analysis,” JAMA Network Open, vol. 4, no. 12, pp. e2137257–e2137257, 2021.

[46] D. P. Oran and E. J. Topol, “Prevalence of asymptomatic SARS-CoV-2 infection: a narrative review,” Annals of Internal Medicine, vol. 173, no. 5, pp. 362–367, 2020.

[47] M. Yanes-Lane, N. Winters, F. Fregonese, M. Bastos, S. Perlman-Arrow, J. R. Campbell, and D. Menzies, “Proportion of asymptomatic infection among COVID-19 positive persons and their transmission potential: A systematic review and meta-analysis,” PloS One, vol. 15, no. 11, p. e0241536, 2020.

[48] N. M. Ferguson, D. Laydon, G. Nedjati-Gilani, N. Imai, K. Ainslie, M. Baguelin, S. Bhatia, A. Boonyasiri, Z. Cucunuba, G. Cuomo-Dannenburg, A. Dighe, I. Dorigatti, H. Fu, K. Gaythorpe, W. Green, A. Hamlet, W. Hinsley, L. C. Okell, S. van--Elsland, H. Thompson, R. Verity, E. Volz, H. Wang, Y. Wang, P. G. Walker, C. Walters, P. Winskill, C. Whittaker, C. A. Donnelly, S. Riley, and A. C. Ghani, “Report 9: Impact of non-pharmaceutical interventions (NPIs) to reduce COVID19 mortality and healthcare demand,” tech. rep., Imperial College London, Mar. 2020.

[49] Y. Niu, J. Rui, Q. Wang, W. Zhang, Z. Chen, F. Xie, Z. Zhao, S. Lin, Y. Zhu, Y. Wang, et al., “Containing the transmission of COVID-19: a modeling study in 160 countries,” Frontiers in Medicine, vol. 8, p. 701836, 2021.

[50] C. N. Ngonghala, J. R. Knitter, L. Marinacci, M. H. Bonds, and A. B. Gumel, “Assessing the impact of widespread respirator use in curtailing COVID-19 transmission in the USA,” Royal Society Open Science, vol. 8, no. 9, p. 210699, 2021.

[51] C. N. Ngonghala, E. Iboi, S. Eikenberry, M. Scotch, C. R. MacIntyre, M. H. Bonds, and A. B. Gumel, “Mathematical assessment of the impact of non-pharmaceutical interventions on curtailing the 2019 novel coronavirus,” Mathematical Biosciences, vol. 325, p. 108364, 2020.

[52] “COVID states project.” https://lazerlab.shinyapps.io/Behaviors_During_COVID/. Accessed December 9, 2023.

[53] CSSE at Johns Hopkins University, “CSSE GIS and Data COVID-19.” https://github.com/CSSEGISandData/COVID-19, 2020.

[54] M. Martcheva, An Introduction to Mathematical Epidemiology, vol. 61. Springer, 2015.

[55] S. M. Kissler, C. Tedijanto, E. Goldstein, Y. H. Grad, and M. Lipsitch, “Projecting the transmission dynamics of SARS-CoV-2 through the postpandemic period,” Science, vol. 368, pp. 860–868, May 2020. Publisher: American Association for the Advancement of Science Section: Report.

[56] R. Subramanian, Q. He, and M. Pascual, “Quantifying asymptomatic infection and transmission of COVID-19 in New York City using observed cases, serology, and testing capacity,” Proceedings of the National Academy of Sciences, vol. 118, no. 9, p. e2019716118, 2021.

[57] S. M. Moghadas, M. C. Fitzpatrick, P. Sah, A. Pandey, A. Shoukat, B. H. Singer, and A. P. Galvani, “The implications of silent transmission for the control of COVID-19 outbreaks,” Proceedings of the National Academy of Sciences, vol. 117, no. 30, pp. 17513–17515, 2020.

[58] L. C. Tindale, J. E. Stockdale, M. Coombe, E. S. Garlock, W. Y. V. Lau, M. Saraswat, L. Zhang, D. Chen, J. Wallinga, and C. Colijn, “Evidence for transmission of COVID-19 prior to symptom onset,” Elife, vol. 9, p. e57149, 2020.

[59] A. A. King, M. Domenech de Celles, F. M. Magpantay, and P. Rohani, “Avoidable errors in the modelling of outbreaks of emerging pathogens, with special reference to Ebola,” Proceedings of the Royal Society B: Biological Sciences, vol. 282, no. 1806, p. 20150347, 2015.

[60] B. Ratner, “The correlation coefficient: Its values range between+ 1/-1, or do they?,” Journal of Targeting, Measurement and Analysis for Marketing, vol. 17, no. 2, pp. 139–142, 2009.

[61] V. Lakshmikantham, S. Leela, and A. A. Martynyuk, Stability Analysis of Nonlinear Systems. Springer, 1989.

[62] P. van--den--Driessche and J. Watmough, “Reproduction numbers and sub-threshold endemic equilibria for compartmental models of disease transmission,” Mathematical Biosciences, vol. 180, no. 1-2, pp. 29–48, 2002.

[63] O. Diekmann, J. A. P. Heesterbeek, and J. A. Metz, “On the definition and the computation of the basic reproduction ratio _0_ in models for infectious diseases in heterogeneous populations,” Journal of Mathematical Biology, vol. 28, no. 4, pp. 365–382, 1990.

[64] A. B. Gumel, E. A. Iboi, C. N. Ngonghala, and E. H. Elbasha, “A primer on using mathematics to understand COVID-19 dynamics: Modeling, analysis and simulations,” Infectious Disease Modelling, vol. 6, pp. 148–168, 2021.

[65] A. Mallela, J. Neumann, E. F. Miller, Y. Chen, R. G. Posner, Y. T. Lin, and W. S. Hlavacek, “Bayesian inference of state-level COVID-19 basic reproduction numbers across the United States,” Viruses, vol. 14, no. 1, p. 157, 2022.

[66] W. Yu, Y. Guo, S. Zhang, Y. Kong, Z. Shen, and J. Zhang, “Proportion of asymptomatic infection and nonsevere disease caused by SARS-CoV-2 Omicron variant: A systematic review and analysis,” Journal of Medical Virology, vol. 94, no. 12, pp. 5790–5801, 2022.

[67] P. Sah, M. C. Fitzpatrick, C. F. Zimmer, E. Abdollahi, L. Juden-Kelly, S. M. Moghadas, B. H. Singer, and A. P. Galvani, “Asymptomatic SARS-CoV-2 infection: A systematic review and meta-analysis,” Proceedings of the National Academy of Sciences, vol. 118, no. 34, p. e2109229118, 2021.

[68] D. Han, R. Li, Y. Han, R. Zhang, and J. Li, “Covid-19: Insight into the asymptomatic sars-cov-2 infection and transmission,” International Journal of Biological Sciences, vol. 16, no. 15, p. 2803, 2020.

[69] M. A. Johansson, T. M. Quandelacy, S. Kada, P. V. Prasad, M. Steele, J. T. Brooks, R. B. Slayton, M. Biggerstaff, and J. C. Butler, “SARS-CoV-2 transmission from people without COVID-19 symptoms,” JAMA Network Open, vol. 4, no. 1, pp. e2035057–e2035057, 2021.

[70] A. Chang, M. Sainato, N. Lakhani, R. Kamal, and A. Uteuova, “The pandemic exposed the human cost of the meat-packing industry’s power:’it’s enormously frightening.’.” https://www.theguardian.com/environment/2021/nov/16/meatpacking-industry-covid-outbreaks-workers, 2021. Accessed January 1, 2024.

[71] A. Stewart, I. Kottasová, and A. Khaliq, “Why meat processing plants have become COVID-19 hotbeds,” CNN, June, vol. 27, 2020. Accessed January 1, 2024.

[72] D. Lazer, M. Santillana, R. Perlis, A. Quintana, K. Ognyanova, J. Green, M. Baum, M. D. Simonson, A. Uslu, H. Chwe, et al., “The covid states project# 26: Trajectory of covid-19-related behaviors,” 2021.

[73] T. Chai and R. R. Draxler, “Root mean square error (RMSE) or mean absolute error (MAE)?–arguments against avoiding RMSE in the literature,” Geoscientific Model Development, vol. 7, no. 3, pp. 1247–1250, 2014.

[74] T. O. Hodson, “Root-mean-square error (RMSE) or mean absolute error (MAE): When to use them or not,” Geoscientific Model Development, vol. 15, no. 14, pp. 5481–5487, 2022.

[75] L. R. Kolozsvári, T. Bérczes, A. Hajdu, R. Gesztelyi, A. Tiba, I. Varga, B. Ala’a, G. J. Szőllősi, S. Harsányi, S. Garbóczy, et al., “Predicting the epidemic curve of the coronavirus (SARS-CoV-2) disease (COVID-19) using artificial intelligence: An application on the first and second waves,” Informatics in Medicine Unlocked, vol. 25, p. 100691, 2021.

